# Characterizing SARS-CoV-2 transcription of subgenomic and genomic RNAs during early human infection using multiplexed ddPCR

**DOI:** 10.1101/2022.02.18.22271199

**Authors:** Hyon S. Hwang, Che-Min Lo, Michael Murphy, Tanner Grudda, Nicholas Gallagher, Chun Huai Luo, Matthew L. Robinson, Agha Mirza, Madison Conte, Abigail Conte, Ruifeng Zhou, Christopher B. Brooke, Andrew Pekosz, Heba H. Mostafa, Yukari C. Manabe, Chloe L. Thio, Ashwin Balagopal

**Author notes:** share first co-authorship (Hyon S. Hwang, Che-Min Lo). share last co-authorship (Chloe L. Thio, Ashwin Balagopal). **Corresponding authors:** Ashwin Balagopal or Chloe L. Thio, 855 N. Wolfe Street, Room 535 (AB) or Room 533 (CLT), Baltimore, Maryland, 21205, USA.; (AB).; (CLT).

## Abstract

Control of SARS-CoV-2 (SCV-2) transmission is a major priority that requires understanding SCV-2 replication dynamics. We developed and validated novel droplet digital PCR (ddPCR) assays to quantify SCV-2 subgenomic RNAs (sgRNAs), which are only produced during active viral replication, and discriminate them from full-length genomic RNAs (gRNAs) in a multiplexed format. We applied this multiplex ddPCR assay to 144 cross-sectional nasopharyngeal samples. sgRNAs were quantifiable across a range of qPCR cycle threshold (Ct) values and correlated with Ct values. The ratio of sgRNA:gRNA was remarkably stable across a wide range of Ct values, whereas adjusted amounts of N sgRNA to a human housekeeping gene declined with higher Ct values. Interestingly, adjusted sgRNA and gRNA amounts were quantifiable in culture-negative samples, although levels were significantly lower than in culture-positive samples. Longitudinal daily testing of 6 persons for up to 14 days revealed that sgRNA is concordant with culture results during the first week of infection but may be discordant with culture later in infection. Further, sgRNA:gRNA is constant during infection despite changes in viral culture. These data indicate stable viral transcription during infection. More work is needed to understand why cultures are negative despite persistence of viral RNAs.

## Introduction

Severe acute respiratory syndrome coronavirus 2 (SCV-2) is the etiologic agent of COVID-19, which is primarily a respiratory tract infection with a spectrum of clinical outcomes ranging from mild disease to Acute Respiratory Distress Syndrome (ARDS) and death. In the time since its discovery, the SCV-2 pandemic has resulted in over 400 million cases and over 5 million deaths worldwide (1). In the United States, there are over 77 million cases and over 900,000 deaths. Remarkable progress has been made with SCV-2 vaccines, but emerging variants with higher infectiousness complicate our ability to achieve herd immunity; thus, understanding SCV-2 replication dynamics and infectiousness in early infection are critical to controlling the pandemic.

Viral abundance of SCV-2 from nasal swabs has been qualitatively estimated by using the cycle threshold (Ct) values produced by the reverse transcription PCR (RT-PCR), but there is concern that Ct values of these samples may not always correspond with their infectiousness (2, 3). Additionally, there may be variant-specific differences in viral infectiousness in samples that have similar Ct values, irrespective of vaccination status (4). In contrast, virus culture, the current gold standard method for determining the presence of infectious SCV-2 from a nasal swab sample, has limited sensitivity, requires a biosafety level 3 (BSL-3) containment, and requires highly-skilled technicians to perform. Wolfel et al. reported that culture was only positive when viral RNA levels were ≥ 10^6^ copies/mL, and only routinely detectable at ≥ 10^8^ copies/mL (5). The limited sensitivity of culture is further underscored in a study by Arons et al. where culture was positive from 10 of 16 specimens from people with typical COVID-19 symptoms with detectable RNA by conventional PCR. In addition, culture was only positive in 21 of 30 people with atypical or no symptoms who had detectable SCV-2 RNA by RT-qPCR (6). In that study, culture was negative even in individuals with abundant viral RNA as evidenced by low Ct values. The true duration that infectious virus is present is unknown in most people, although it has been modeled (7). It is now recommended that Ct values from conventional RT-PCR not be used as a surrogate for active viral replication, chiefly due to a poor understanding of whether certain Ct values represent infectious virus or simply the detection of inert viral RNAs, and also due to methodological differences between laboratories (8, 9). This uncertainty raises the question of how best to use Ct values in a clinical setting during the natural course of infection.

SCV-2 is a positive strand RNA virus that gains entry into host cells by the binding of its spike (S) protein to the human ACE2 (hACE2) and TMPRSS2 receptors. In the cell, the ∼30kb positive-strand genomic RNA (gRNA) is translated to produce the polypeptides ORF1a and ORF1b that are proteolytically cleaved into non-structural proteins including the RNA dependent RNA polymerase (RdRp) (10). The RdRp uses the positive-strand gRNA template to transcribe full-length negative-strand intermediates and a set of negative-strand subgenomic RNAs (sgRNA). The RdRp then uses the full-length negative-strand intermediates as templates to produce full-length positive-strand gRNA for progeny viruses while using the negative-strand sgRNAs as templates for positive-strand sgRNA production that are translated to structural proteins nucleocapsid (N), spike (S), envelope (E), and membrane (M) as well as several accessory proteins. sgRNA is chiefly produced in cells with active replication and is not packaged into virions. Thus, viral transcription is a critical step in virus replication, and transcription of sgRNAs and gRNAs indicates active cellular processes that are consistent with ongoing viral replication. However, an extant question is whether the presence of sgRNA in a clinical sample signifies the potential for infectiousness and transmissibility to susceptible people.

Since the production of SCV-2 sgRNA represents ongoing viral transcription, an E sgRNA assay developed by Wolfel et al. has been utilized commonly to study replication dynamics (5, 11–17). Few studies showed that sgRNA decreased with respect to the amount of gRNA over time, but they did not report the duration of sgRNA shedding (11, 15). In another study, the investigators found a constant ratio of sgRNA to gRNA in hospitalized patients with respect to total SCV-2 RNA (12). However, their assay did not directly measure gRNA, and was not multiplexed with sgRNA as also seen in most studies, so inherent differences in their sgRNA and gRNA assays could exist. Truong et al. performed viral culture to confirm infectivity and reported a strong correlation between sgRNA quantity and viral culture data, suggesting that sgRNA may serve as a molecular marker for viral infectivity (17).

Due to limited yield of viral RNA from swab samples and the presence of potential inhibitors in the collection medium, droplet digital PCR is ideal for sensitive, reliable, and simultaneous quantitation of sgRNA and gRNA. Few studies have used ddPCR to examine viral kinetics on limited number of subjects or time points per participant (12, 16, 18, 19); therefore, more extensive cross-sectional and longitudinal sample analyses are desired for full characterization of viral kinetics. To address these knowledge gaps and to link active SCV-2 transcription with infectiousness, we developed a multiplex ddPCR assay to quantify SCV-2 sgRNA and gRNA simultaneously and applied this assay to cross-sectional and longitudinal samples obtained from participants with recent COVID-19 who also had viral culture performed.

## Results

### Validating a multiplex ddPCR assay for multiple SCV-2 targets

SCV-2 gRNA and sgRNAs share a common 5’ leader sequence of ∼70 nucleotides that is fused to the body sequence: individual sgRNAs are transcribed when the viral replication-transcription complex recombines the 5’ leader sequence with downstream gene targets. We designed ddPCR primers and probes that exploited this common 5’ leader sequence to specifically amplify the N, S, and E sgRNAs (Supplemental Figure 1; Supplemental Table). We tested the performance of sgRNA and gRNA ddPCR assays against known quantities of synthesized SCV-2 gene fragments for each target individually and in multiplex assays, followed by validation with clinical sample RNA extracts from clinical samples with varying SCV-2 qPCR Ct levels, along with five negative control samples from healthy donors. Individually, all ddPCR sgRNA assays (N, E, and S) and the gRNA assay had similar efficiencies. We incorporated the N sgRNA assay into a multiplexed assay with gRNA for subsequent testing since N sgRNA was more abundant than E and S sgRNAs in our clinical samples (data not shown) and has been described as the most abundantly expressed sgRNA (10). We found that each N sgRNA and gRNA ddPCR assay performed well individually with a lower limit of detection (LLOD) of 1.1 absolute copies for each target individually and in multiplex assays. The lower limits of quantitation (LLOQ) were 3.2 copies for gRNA, 3.5 for N sgRNA, and 4.5 for both gRNA and N sgRNA in the multiplex assay (Supplemental Figure 2). When mixing distinct targets, we did not observe false identification of sgRNA or gRNA target as the other, demonstrating that the assays were specific and exclusive for each target individually. To confirm specificity of each assay, we performed multiplex ddPCR for N sgRNA and gRNA on nasal swabs from five healthy volunteers who had no symptoms and no known exposures within 14 days of sampling: we did not detect any evidence of N sgRNA nor gRNA in these samples.

### Cross-sectional testing

We applied our multiplex ddPCR assay to 144 RNA extracts from participants in an anonymized cross-sectional cohort with confirmed SCV-2 infection. The median (IQR) N sgRNA amounts were 2.1 x 10^6^ (2.3 x 10^5^-2.9 x10^7^) copies/mL and of gRNA were 6.4 x 10^5^ (4.6 x 10^4^-1.4 x 10^7^) copies/mL. Normalized to RPLP1 quantities in the same samples (see methods), the median (IQR) amounts of sgRNA and gRNA were 5.8 (0.4-62.8) and 1.4 (0.1-22.4), respectively. The median (range) Ct values for total SCV-2 RNA was 20.7 (17.3-24.7). Normalized quantities of sgRNA and gRNA correlated well with total amounts of SCV-2 RNA (Ct values) (r=0.76, p<0.05; Figure 1).

**Figure 1.**
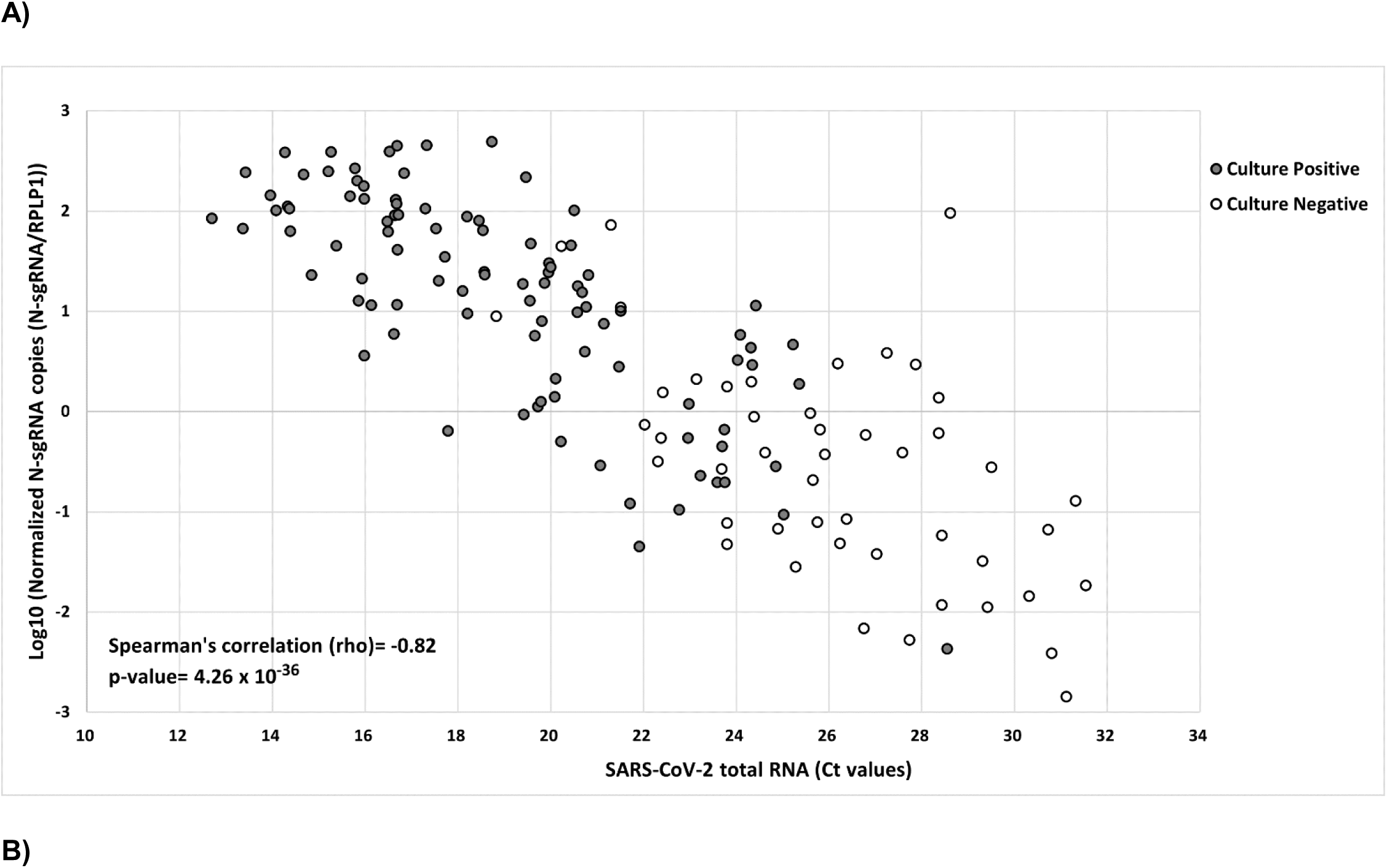
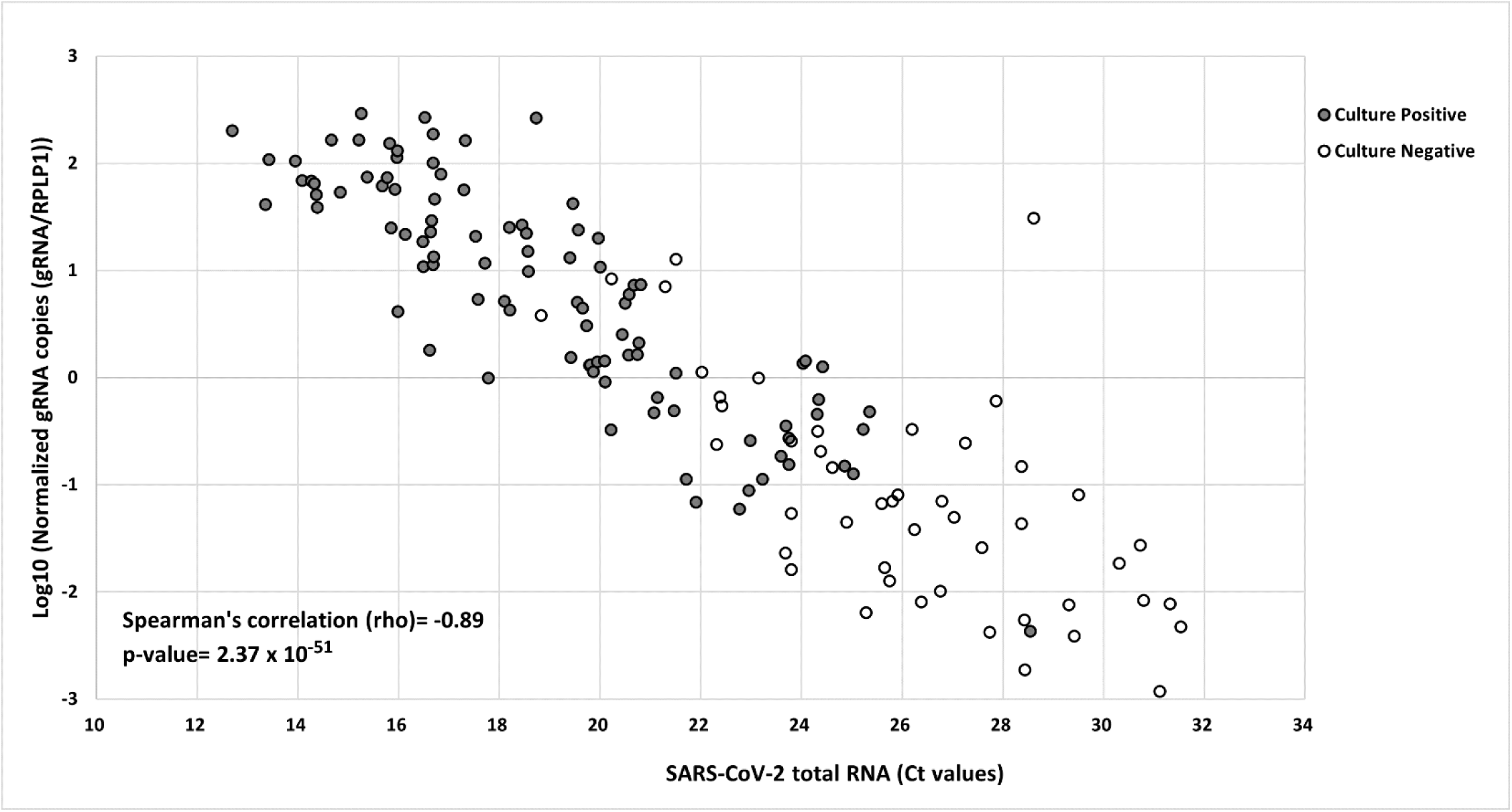
ddPCR of adjusted N sgRNA and gRNA values are correlated with qPCR amounts. Multiplex ddPCR for A) N sgRNA and B) gRNA was performed on NP swabs taken from 144 people in the JHCVL cohort. All values were adjusted for the amount of host mRNA RPLP1 in the same sample, also measured by ddPCR. On the x-axis is shown the amount of total SCV-2 RNA quantified in the same sample, as measured by qPCR. On the y-axis is shown the adjusted ddPCR results, log_10_ transformed. Culture results in the same samples is depicted as open (culture-negative) and closed (culture-positive) circles. Spearman’s correlation coefficients and corresponding p-values were calculated and displayed.

Viral culture had been performed from an aliquot of the same samples in each person (see methods) and resulted as positive (n=97) or negative (n=47) (Figure 2). Samples were then categorized based on qPCR Ct value and compared to the normalized amount of sgRNA/RPLP1 (Figure 3). Notably, 41 samples with Ct values < 18 were all culture-positive and 9 samples with Ct values ≥ 29 were all culture-negative, but there was marked heterogeneity in culture results between these Ct values (Supplemental Figure 3). Thus, Ct values alone were inadequate to predict culture positivity. Importantly, normalized N sgRNA and gRNA values were significantly different between culture-positive and culture-negative samples in intermediate range of Ct values (p= 4.18 x 10^-4^ and 7.95 x 10^-6^, respectively) (Figure 3). Surprisingly, although there were small differences in raw sgRNA or gRNA values over the same intermediate range of Ct values (Supplemental Figure 3), raw values were insufficient to discriminate between culture-positive and culture-negative samples. However, it should also be noted that in this intermediate range, the mean Ct values for the culture-positive samples were significantly lower than the culture-negative samples (21.3 vs 25.1, p= 2.39 x 10^-8^). Taken together, samples that were culture-positive had significantly higher normalized N sgRNA (95% CI 50.2-95.9) amounts than samples that were culture-negative (95% CI 0.2-10.9). Also, higher amounts of normalized gRNA (95% CI 25.7-51.3) were demonstrated in culture-positive samples than in culture-negative (95% CI 0.01-2.9) (Figure 2). These results strongly suggest that when the proportion of total RNA that was viral is high, samples were more likely to be culture-positive, whereas when the proportion of total RNA that was viral is low, samples were more likely to be culture-negative. As a separate marker of abundance, we categorized culture-positive samples based on when they turned positive in vitro, which was ascertained between 2-5 days post inoculation (dpi). Samples that turned culture-positive on 2 dpi had consistently more total SCV-2 RNA (measured by qPCR), normalized sgRNA (ddPCR), and normalized gRNA (ddPCR) compared to samples that turned culture-positive at later dates (Supplemental Figure 4). These results are consistent with the hypothesis that the normalized sgRNA and gRNA values are indicative of the fraction of viral RNAs that are replication competent in culture.

**Figure 2.**
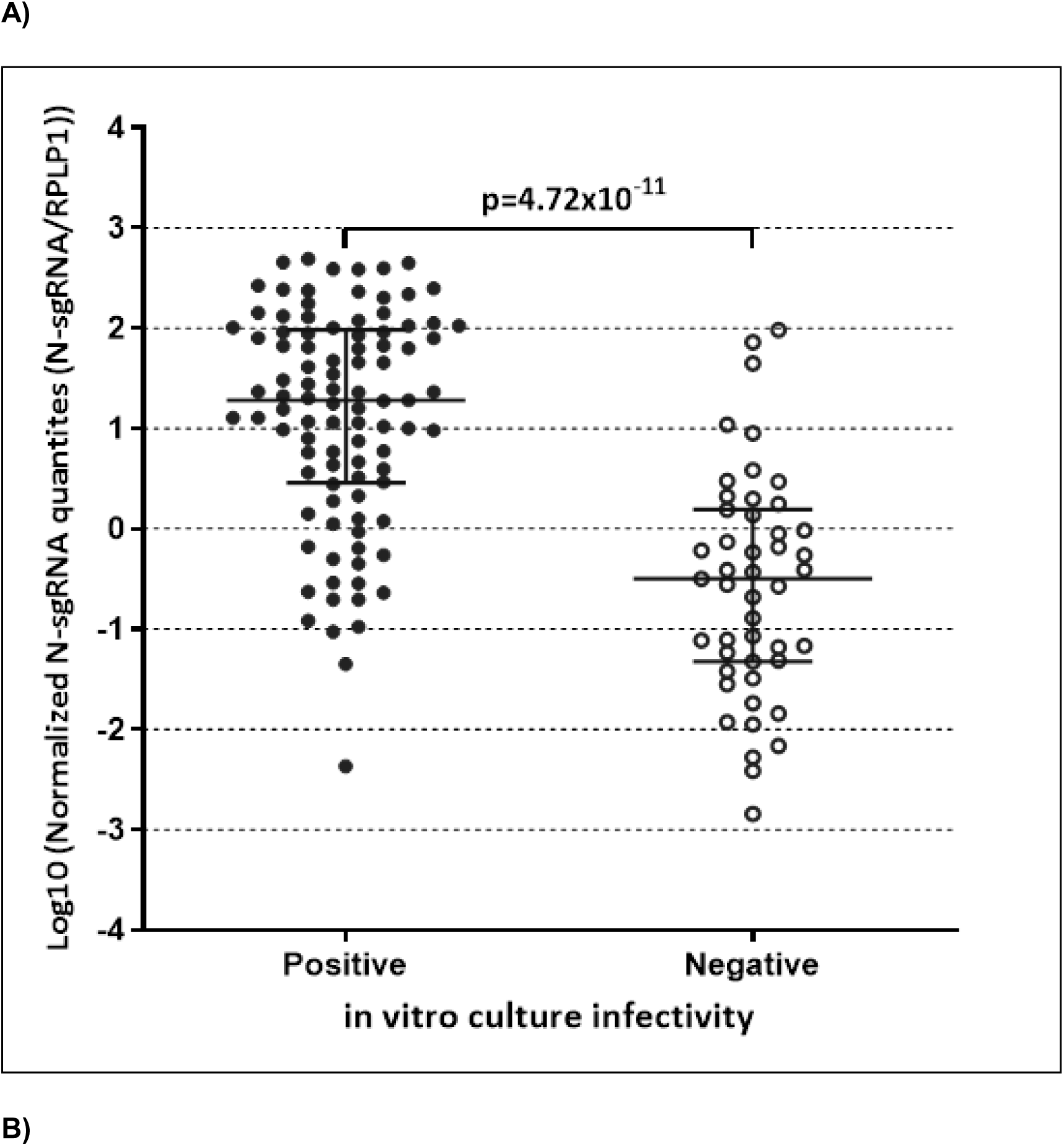
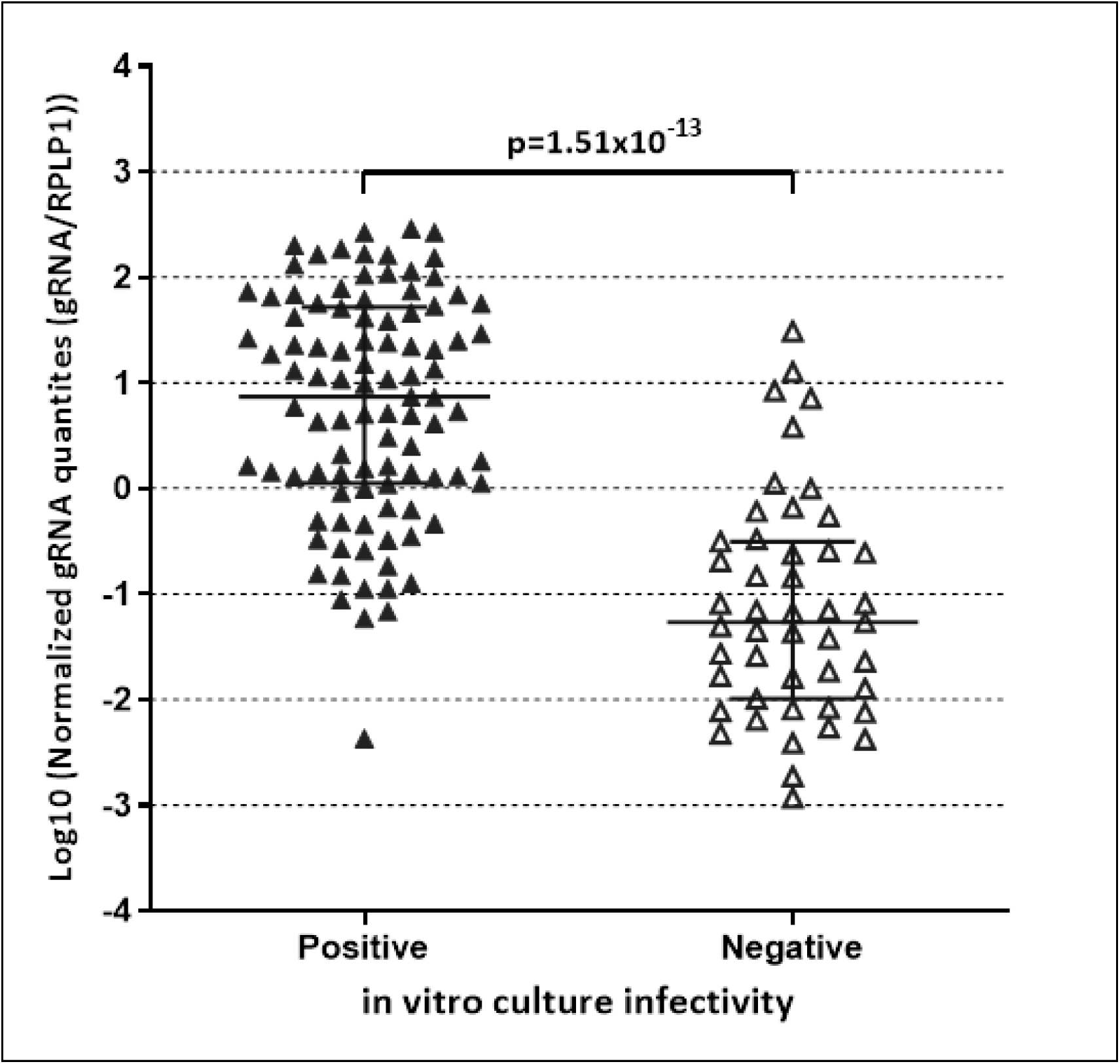
Adjusted N sgRNA and gRNA levels stratified by virus culture results. Multiplex ddPCR results for A) N sgRNA and B) gRNA are shown for 144 cross-sectional JHCVL samples that also had virus culture performed. Shown are log_10_ values, adjusted for RPLP1 mRNA in the same samples. Wilcoxon rank sum tests were used to compare adjusted quantities in culture-positive and culture-negative samples.

**Figure 3.**
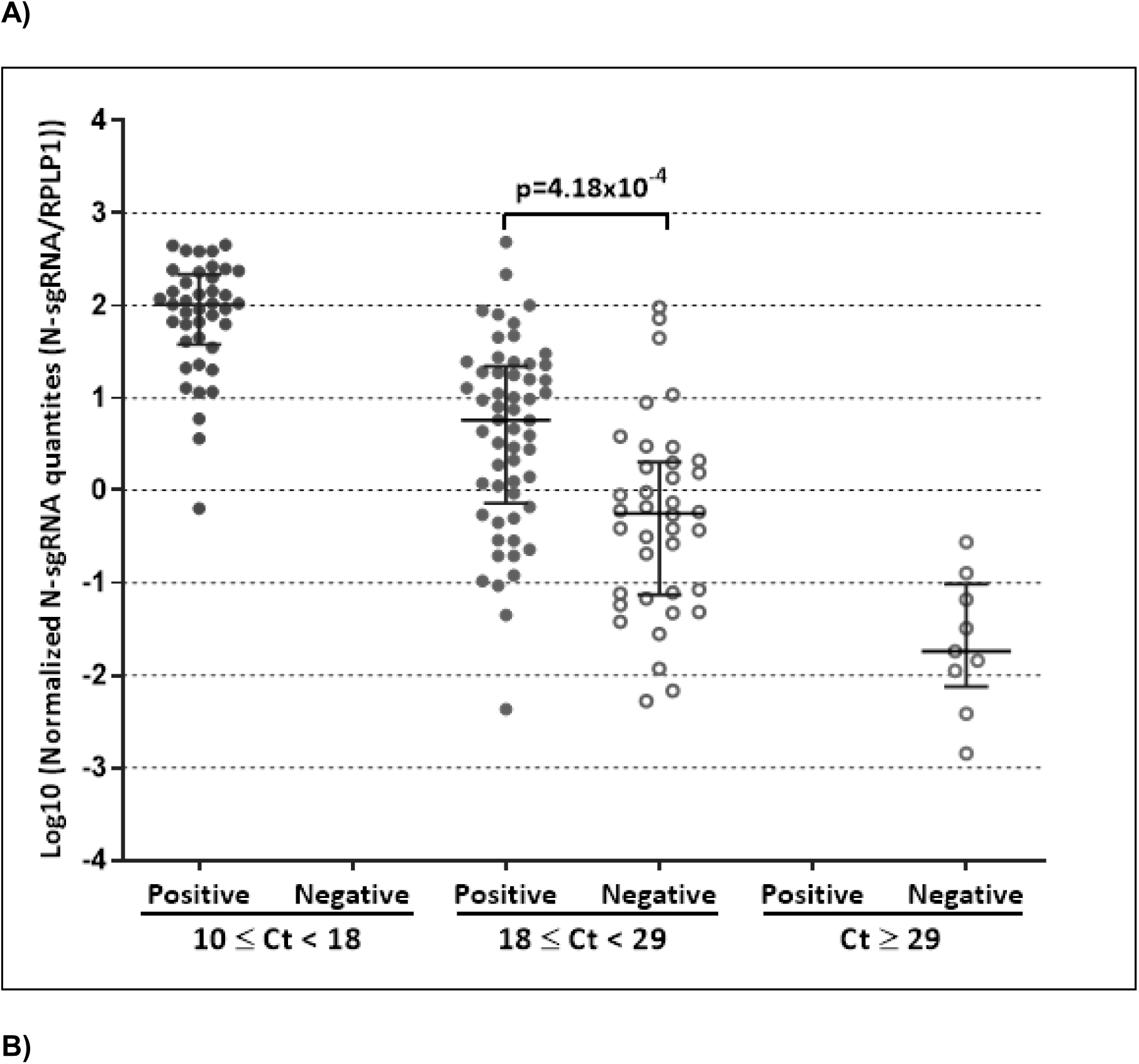
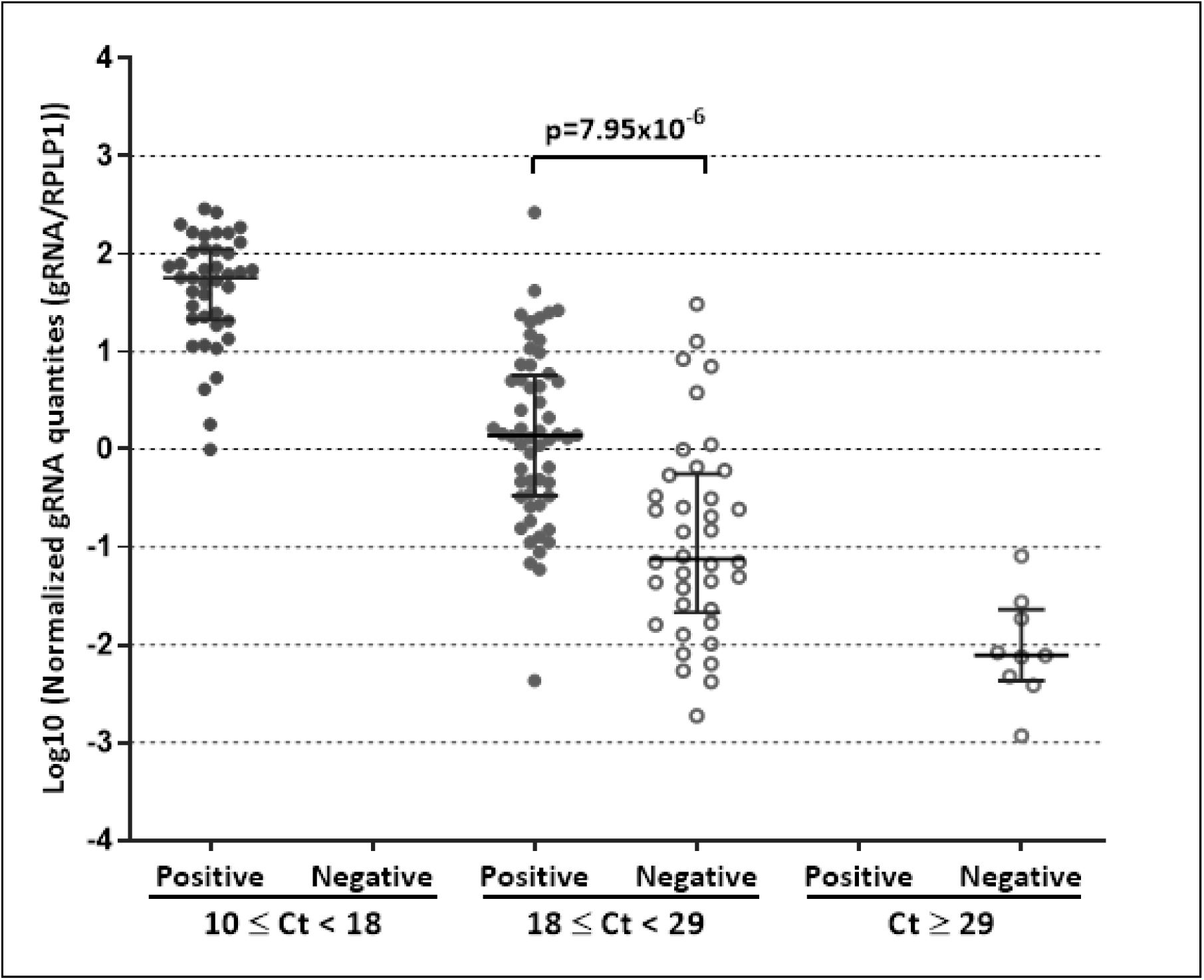
Adjusted N sgRNA quantities stratified by qPCR and viral culture results. Log_10_ adjusted N sgRNA values are shown for samples stratified by total SCV-2 RNA abundance, as measured by qPCR. Quantities in culture-positive (closed circles) and culture-negative (open circles) samples are shown. Kruskall-Wallis tests were used to compare adjusted N sgRNA quantities in culture-positive versus negative samples with intermediate Ct values of 18-29.

### Longitudinal testing

To address sgRNA and gRNA production during the course of infection, we tested samples taken from nasal swabs that were obtained from 27 persons in the UIUC cohort at two time points (see methods). The first time point was collected a median (IQR) 1 (1-1) days after symptom onset and the second time point was collected a median (IQR) 5 (4-7) days after symptom onset. When these sgRNA and gRNA measurements were overlaid on daily Ct values, annotated by culture result, the normalized sgRNA and gRNA generally tracked with the Ct values (Figure 4; Supplemental Figure 5). 24/27 first time point samples were culture-positive while 13/27 second time point samples were culture-positive. Of the 13 culture-positives at the second time point, only one was negative at the first time point (Supplemental Figure 5) while the other 12 were positive at both time points. Among persons for whom the first time point samples were culture-positive, 12/24 (50%) second time point samples were culture-negative. Unexpectedly, in 5 of these 12 that went from culture-positive to negative, the normalized sgRNA and gRNA values were higher at the culture-negative time point than at the culture-positive time point. The lack of correspondence between sgRNA measurements and culture positivity was largely explained by the viral kinetics as measured by qPCR of total SCV-2 RNA: nearly all culture-positive-negative pairs that had increases in sgRNA or gRNA levels in late samples occurred after the peak in total SCV-2 RNA. Thus, peak total SCV-2 RNA abundance as measured by qPCR demarcated culture-positive versus -negative results irrespective of total, sgRNA, or gRNA abundance. In 6/27 persons (including the 5 individuals with discordance between sgRNA and culture results) who had intensive (daily) longitudinal sampling performed, we confirmed that normalized sgRNA and gRNA amounts measured by ddPCR tracked closely with total RNA amounts measured by qPCR (Figure 5).

**Figure 4.**
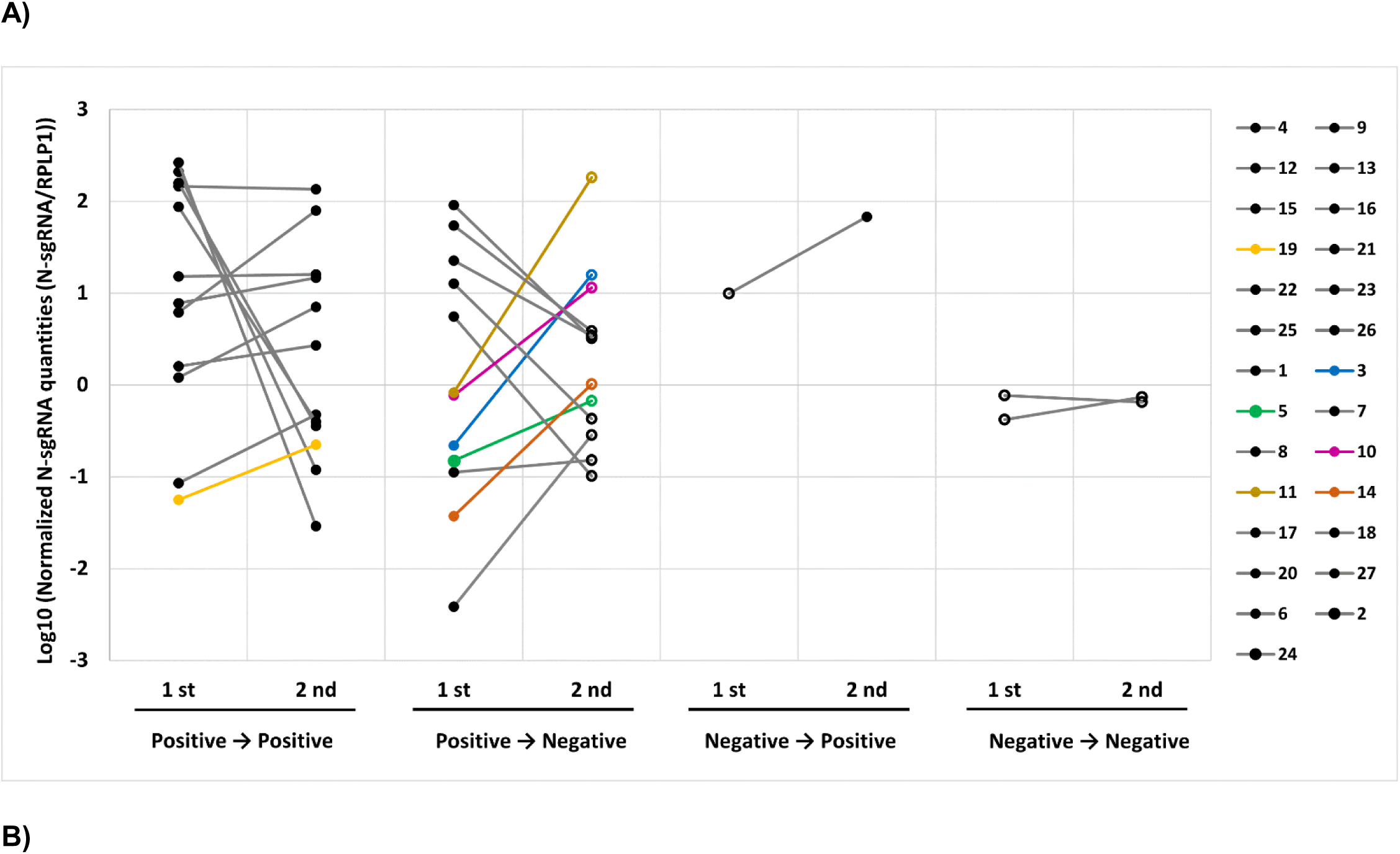
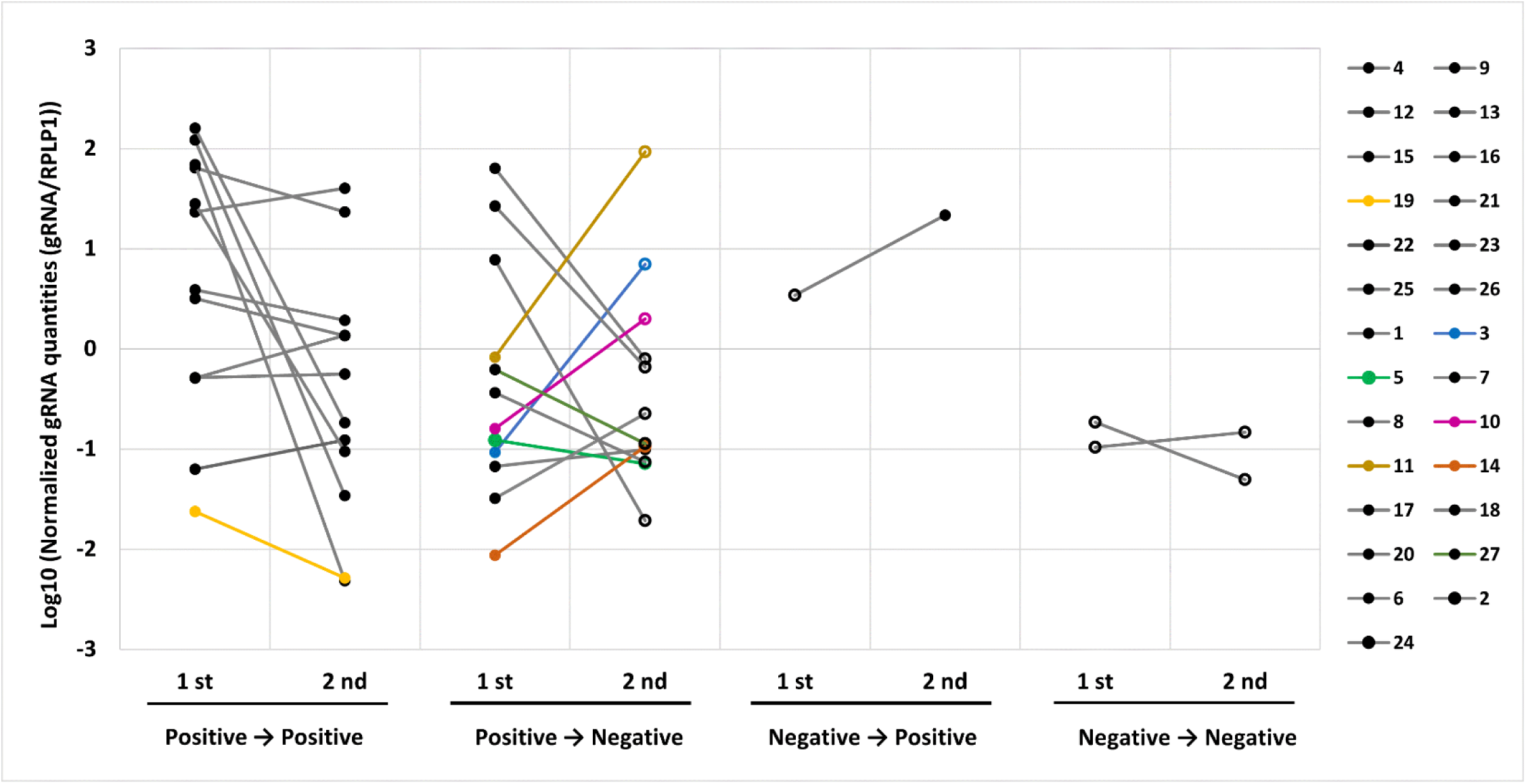
Early and late adjusted N sgRNA and gRNA quantities and virus culture results. Multiplex ddPCR for A) N sgRNA and B) gRNA was performed on two samples from 27 UIUC persons with SCV-2 infection, separated by time. Values were adjusted for quantities of RPLP1 mRNA in the same samples. The 27 persons were categorized into four groups based on virus culture results from contemporaneous samples: those that were culture-positive early and late (Pos→Pos), those that were culture-positive early and culture-negative late (Pos→Neg), those that were culture-negative early and culture-positive late (Neg→Pos), and those that were culture-negative at both times (Neg→Neg). Legend and colors indicate participant ID number. Samples in color indicate those with either adjusted sgRNA or gRNA results that were discrepant with culture results and were then examined in greater detail longitudinally.

**Figure 5.**
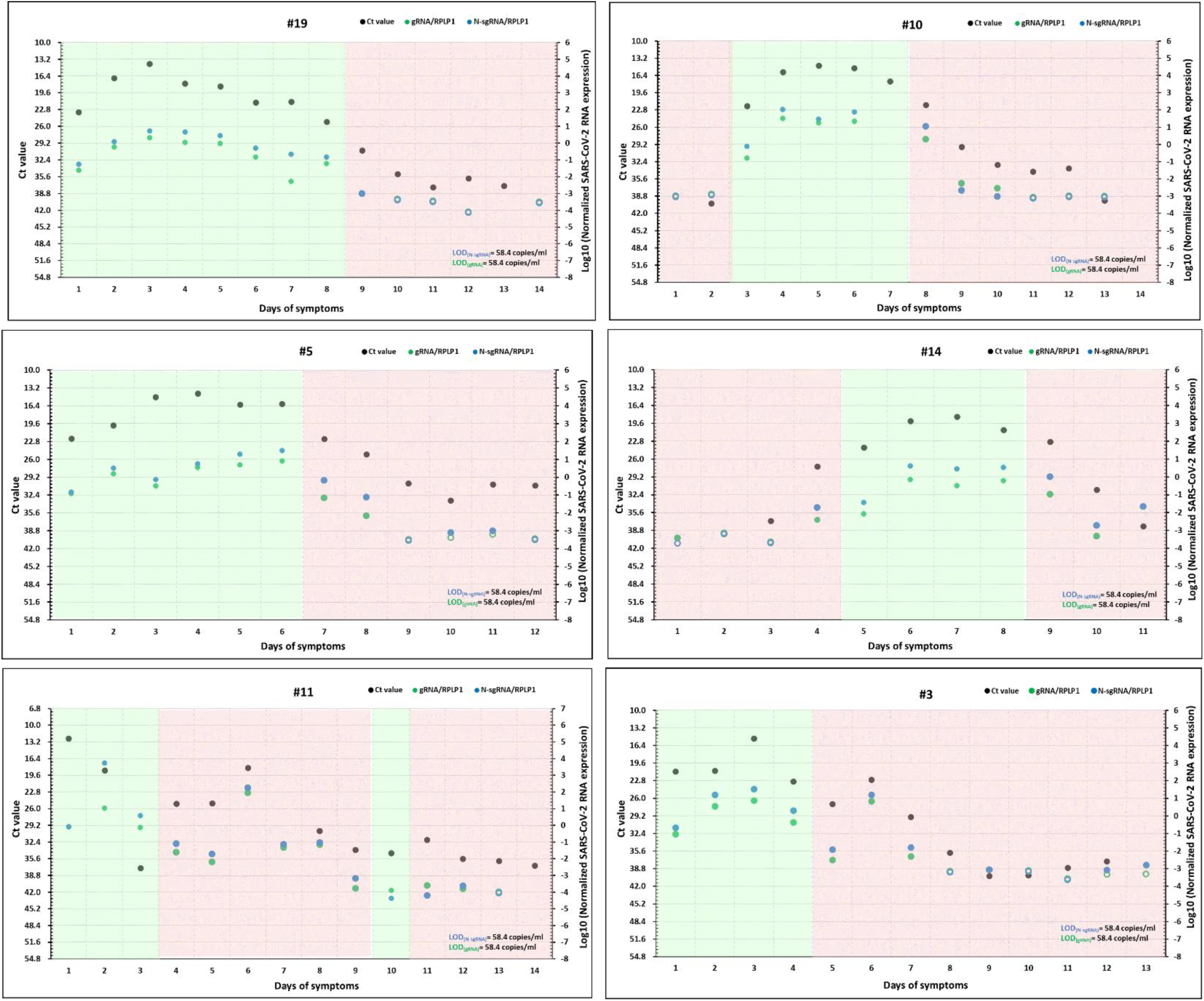
Longitudinal adjusted N sgRNA and gRNA quantities, total SCV-2 RNA, and virus culture results. In 6 persons in the UIUC cohort (identified in Figure 4), daily nasal swabs were tested for multiple RNAs: N sgRNA (by ddPCR), gRNA (by ddPCR), total SCV-2 RNA (by qPCR), and RPLP1 (by ddPCR). Virus culture was also performed on daily samples. Shown are total SCV-2 RNA (Ct values – black points), adjusted log_10_ N sgRNA values (blue points), adjusted log_10_ gRNA values (green points), and virus culture results (pink shading – culture-negative; green shading – culture-positive). Raw values (unadjusted) of N sgRNA and gRNA are shown in Supplemental Figure 5. Right and left y-axes have identical relative scales.

### Testing for differences in transcription of sgRNA vs. gRNA

We next compared the normalized N sgRNA and gRNA quantities obtained from the cross-sectional and longitudinal samples and found a tight correlation (Figure 6). Moreover, the ratio of sgRNA:gRNA was noted to have a narrow range. We then tested the longitudinal samples in UIUC cohort, comparing results in the 21 persons sampled at two time points and the 6 people sampled daily. We interpolated slopes of the sgRNA:gRNA ratios over time for each person and found minimal changes in the ratio during the course of illness (Figure 6; Supplemental Figure 6).

**Figure 6.**
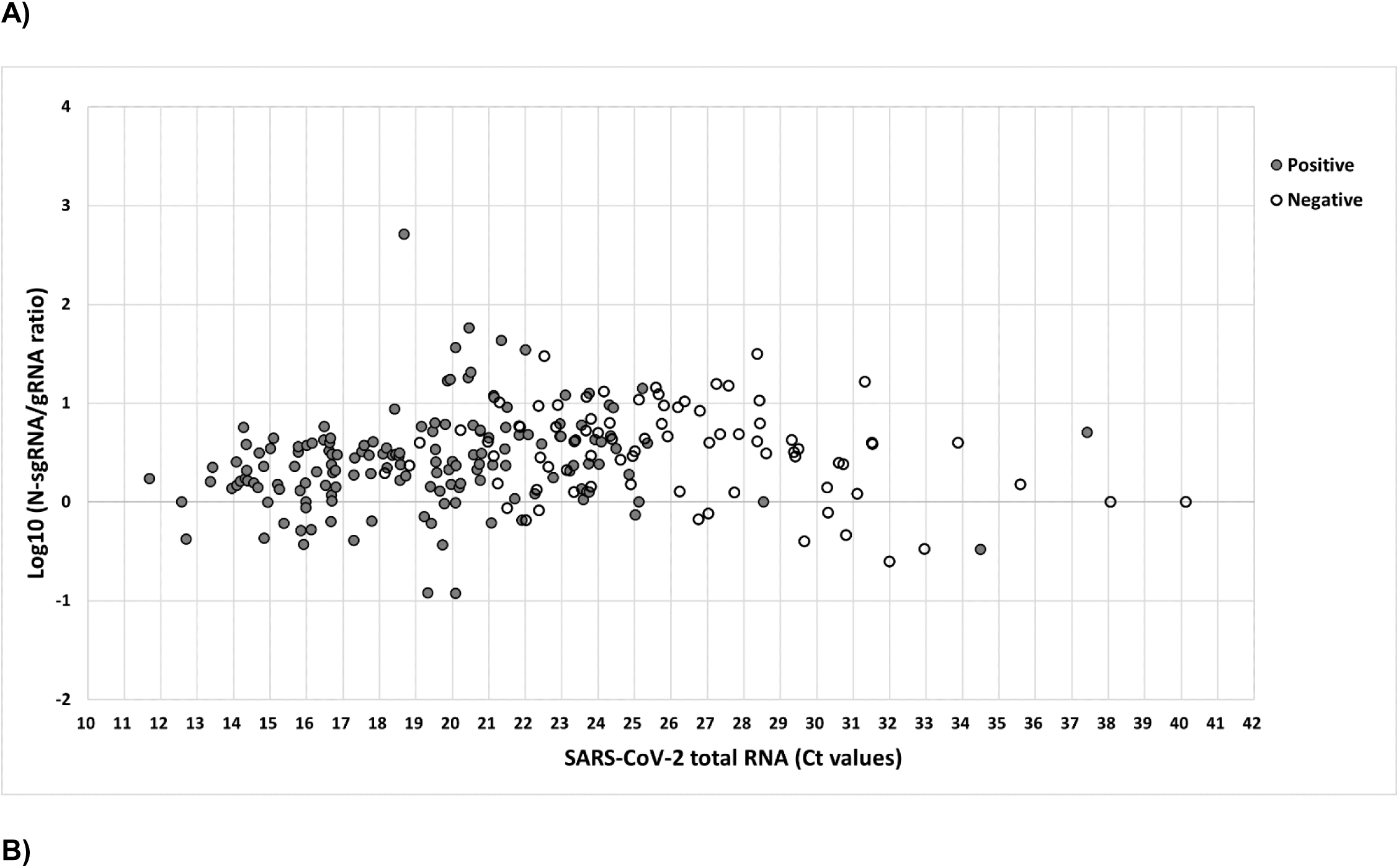
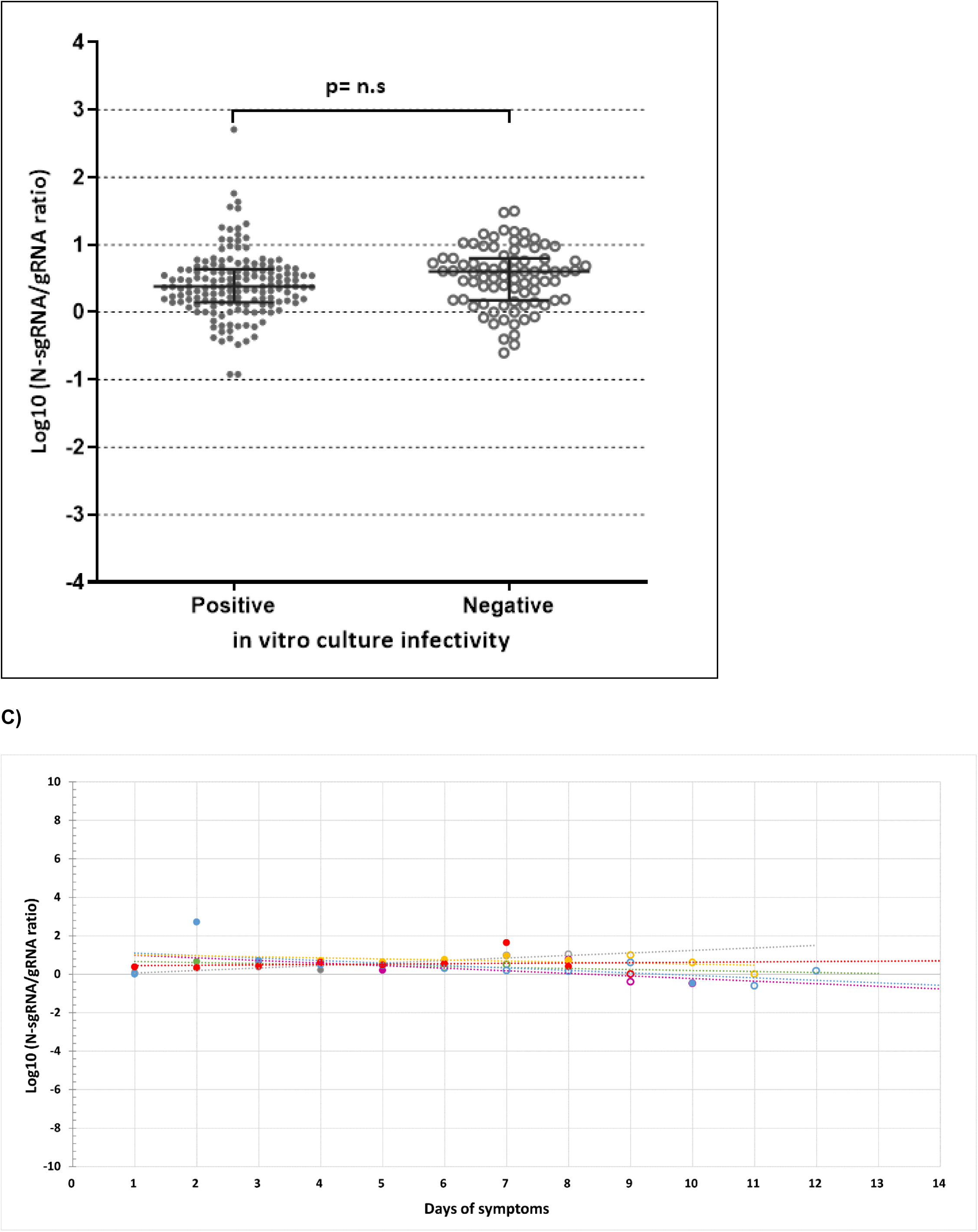
The ratio of N sgRNA:gRNA does not change during SCV-2 infection. Multiple ddPCR results for N sgRNA and gRNA are shown A) across a range of total SCV-2 RNA amounts as indicated by qPCR Ct values and stratified by virus culture results (positive – closed points; negative – open points). Results are from 144 people in the JHCVL cohort (144 samples) and 27 people (2 time points=54 samples) in the UIUC cohort. Displayed are log_10_ N sgRNA:gRNA ratios. B) The N sgRNA:gRNA ratios are not significantly different between virus culture-negative and positive samples. C) In 6/27 persons in the UIUC cohort where daily NP swabs were tested for N sgRNA and gRNA amounts using the multiplex ddPCR assay, longitudinal slopes were generated depicting each individual’s daily N sgRNA:gRNA ratio. Collectively, there were no significant differences in the slopes compared to the null hypothesis that there are no differences in the ratios over time.

## Discussion

This is the first study to use multiplex ddPCR for gRNA and sgRNA to characterize viral transcription, a necessary step in the viral lifecycle, in a large cohort of individuals including careful longitudinal study of six individuals during the first 14 days of infection. We found evidence of abundant viral transcription of sgRNA and gRNA that corresponded closely with total viral RNA as measured by qPCR (Ct values) at all times sampled. Samples that were culture-positive showed higher levels of both total viral RNA and sgRNA; although when these molecular tests were in the mid-range, they did not discriminate culture results as well (Figure 3). The longitudinal samples shed some light on these mid-range samples since those earlier in infection are culture-positive whereas those later in infection tended to be culture-negative. Taken together, these findings suggest that the time course of infection when combined with viral abundance is a better determinant of infectiousness than individual quantities of total, sgRNA, or gRNA, contrary to our original hypothesis. This raises the question as to whether viral transcription that results in the production of infectious virions early is altered over time to favor less infectious virions later during infection. Our data demonstrating a relatively constant ratio of sgRNA:gRNA do not support this hypothesis. Since the sgRNA:gRNA ratios were not substantially different over the course of infection, it is more likely that the SCV-2 specific antibody response that emerges within 14-21 days determines whether an individual is infectious rather than a fixed quantity of viral RNA.

Since qPCR detects gRNA, which may not be indicative of a productive infection, an ongoing question is whether a positive qPCR, especially later in infection, represents shedding of inert virus or infectious virions. In the African green monkey model, gRNA was shown to be highly stable and detectable in the absence of active viral transcription, since it is likely protected from degradation by the viral capsid. However, sgRNAs were degraded quickly and were only detectable during active viral transcription (24). Our data from ddPCR clearly demonstrate that the amount of total viral RNA correlates well with the amount of sgRNA. Thus, a positive qPCR does not simply represent viral shedding, since we found sgRNA produced concurrently. Two other studies also found a high correlation between gRNA and sgRNA but culture was not performed in these studies, limiting the ability to conclude whether samples were infectious (12, 14). Both papers also found that sgRNA can be shed for over 14 days in some individuals.

Although the amount of sgRNA produced was greater in samples that were culture-positive than culture-negative, there were some samples with detectable sgRNA that were culture-negative. The longitudinal samples (UIUC) demonstrated that in some cases culture-negative samples had quantities of sgRNA equivalent to culture-positive samples. This finding cannot be due to different amounts of RNA extract since we normalized for the total amount of RNA by measuring RPLP1 in the same sample. Since sgRNA signify active replication, but are not themselves packaged into virions, one potential explanation for this discrepancy is that infectious virions are present in these samples but are adsorbed to neutralizing antibodies and targeted for reticulo-endothelial clearance (25). In support of this hypothesis, Killerby et al. determined SCV-2 RNA, culture, and antibody kinetics in 14 persons with COVID-19 followed for 6 months after symptom onset (26). They found that viral cultures turned negative soon or immediately after the detection of neutralizing antibodies. However, quantities of SCV-2 RNA were not provided in that report. An alternative explanation is that continued shedding of sgRNA represents continued release of RNAs from long-lived infected cells, such as ciliated cells in the respiratory mucosa and sustentacular cells in the olfactory mucosa which were identified as the major target cell types for SARS-CoV-2 replication in the nasal mucosa (27). Lastly, it is possible that these sgRNAs have deletions or are mutated so that viral proteins needed to produce virions are not translated; however, it seems unlikely that this would be a frequent occurrence, or that proportionally more viral transcripts would be expected to be defective over time. Understanding culture-negative samples that also have detectable sgRNAs is clinically relevant since it is not yet known how much overlap there is between persons who have prolonged shedding of SCV-2 RNA and persons with past acute sequelae of COVID-19 (PASC), which can occur in up to 1 in 3 persons with COVID-19. It is possible that infectious reservoirs of SCV-2 may sustain or contribute to symptoms in PASC. Solidifying whether there is prolonged viral transcription in these individuals would help to disentangle the viral contribution to the PASC phenotype.

It is notable that these culture-negative/sgRNA positive samples tend to be later in the course of infection. Thus, regardless of which of the above explanations is operative, our data support that in the immunocompetent patient, a combination of the time course of infection along with viral RNA abundance may be a better determinant of the infectious status of an individual rather than one individual marker. Our data do not address immunosuppressed patients in whom longer detection of sgRNA may represent prolonged infectiousness.

It is also intriguing that we found consistent sgRNA:gRNA levels in our cross-sectional and longitudinal cohorts and that the longitudinal samples did not show significant changes in the ratio over time. These findings suggest that viral transcription may not be differentially regulated between producing sgRNAs and full-length genomes that are packaged into virions. While this is necessarily true for smaller RNA viruses that transcribe single strand RNAs such as hepatitis C virus (HCV) that dually function as templates for translation into poly-proteins and genomes for packaging into nascent virions, it was unclear previously whether this was also true for SCV-2, which has a comparatively larger RNA genome and evidence for multiple RNA start sites. Although other studies have found relatively stable levels of sgRNA:gRNA over time, they used separate assays to determine the sgRNA and gRNA levels, so those results could be affected by assay efficiency or interassay variability (5, 12). We overcame these by using our multiplex ddPCR assay that measures sgRNA and gRNA simultaneously. Interestingly, whereas we found consistent sgRNA:gRNA measurements, the amount of sgRNA or gRNA as a proportion of total RNA (quantified by RPLP1 values) did seem to vary with symptom onset, and these results corresponded closely with culture results when obtained early after enrollment. Thus, our data suggest that there are complex dynamics of total RNA, total SCV-2 RNA, and sgRNAs that may not be reflected in single-time point measurements of any individual target.

There are several limitations to our study. The sensitivity of RNA detection in NP samples may not reflect the presence of RNA in deeper and more sequestered compartments of the body. However, by using ddPCR to improve our sensitivity by at least an order of magnitude, and by quantifying human RNA using housekeeping genes, we have at least addressed sensitivity in clinically relevant specimens. A related limitation, however, is that we did not perform a comprehensive analysis of human RNAs in clinical samples, which may have revealed which cells were infected or contributing to COVID-19 pathology, as has been done previously (28). With regards to viral culture, it is well-appreciated that there are limits to its sensitivity. Indeed, as we speculate above, culture negativity does not reveal whether virions are replication incompetent or are adsorbed to antibodies and thus neutralized in the culture experiment. Unfortunately, samples for antibody testing were not available, but it is possible that antibody testing may have explained why some culture samples were negative despite deriving from NP swabs that had abundant amounts of sgRNA, gRNA, and total viral RNA. Nevertheless, culture negativity does correspond well with when COVID-19 patients are no longer likely to transmit to others, and thus is still a useful tool for research, if challenging to implement for widespread clinical use. In addition, we note here that while there was considerable homogeneity in our findings in these clinical cohorts, we did not study any people with immunosuppression for whom viral kinetics might be markedly altered. It is also worth noting that because participants in the UIUC cohort received their positive test results prior to enrollment, this may have influenced their report of symptoms. Finally, although a fine point, we note that measurements of sgRNAs largely reflect measurements of entities that reside strictly intracellularly apart from fragmentation, whereas measurements of total SCV-2 and gRNA reflect entities that could be either intracellular or extracellular. Thus, there is a concern that our inferences may be removed from the biological processes that generated the objects of measure.

In closing, we report here that sgRNA and gRNA levels obtained early after COVID-19 symptom onset can distinguish infectious from non-infectious virions. However, culture results appear to be more governed by duration of illness at later time points, which is highly likely to reflect the antiviral effects of an incipient adaptive immune response. Viral transcription and replication may continue unabated during the immune response. The ramifications of continued viral transcription are unclear but there may be overlap between prolonged viral activity and PASC, which could be addressed in future longitudinal studies.

## Methods

### Cross-sectional samples

After optimization of the ddPCR assay (Supplemental Figure 2), we first tested 144 anonymized nasopharyngeal (NP) swab RNA extracts with available SCV-2 qPCR Ct values and virus culture results from the Johns Hopkins Clinical Virology Laboratory (JHCVL). We selected samples that were qPCR positive over a wide range of Ct values: 97 samples that were culture-positive with a Ct range from 12.70 to 28.54 and 47 samples that were culture-negative with a Ct range from 18.83 to 31.53 (4, 20, 21).

### Longitudinal samples

We obtained longitudinal samples from the University of Illinois at Urbana-Champaign (UIUC) cohort that have been extensively characterized for clinical symptoms (self-reported symptom onset), RT-PCR for SCV-2, and virus culture (22, 23). Participants received a positive test prior to enrollment into the cohort. We tested nasal swab RNA extracts from 27 individuals at two time points (54 samples). The Ct range for 37 virus culture-positive samples was 11.69 to 24.49 and for 17 culture-negative samples was 18.14 to 24.16. We specifically chose samples with low and intermediate Ct values to investigate differences that could be observed in virus culture results, therefore excluding high Ct samples that are overwhelmingly culture-negative. To characterize more fully the dynamics of SCV-2 N sgRNA, we selected 6/27 individuals from the UIUC cohort for intensive longitudinal study and tested their daily nasal swab RNA samples for up to 14 days. The individuals selected were based on what appeared to be a discrepancy between virus culture results, qPCR Ct values, gRNA, and N sgRNA ddPCR results from the testing of the two initial time points.

### Negative controls

Five adult volunteers who were asymptomatic, negative for SCV-2, and had not yet had SCV-2 vaccination had nasal swabs obtained and placed into viral transport media. RNA was extracted and subjected to ddPCR assays as negative controls.

### Positive controls for assay validation

We synthesized DNA fragments including 250 bp SCV-2 gene fragments for each of the target N, E, S sgRNAs and gRNA (gBlocks from Integrated DNA Technologies and GenParts from GenScript). Each DNA fragment included the T7 promoter sequence and was in vitro transcribed to produce each targeted RNA fragment using RiboMAX Large Scale RNA Production Systems (Promega; Cat. P1280), followed by quantitation of RNA with Qubit RNA Broad Range Assay Kit (ThermoFisher; Cat. Q10210).

### Nasal swab qPCR, virus culture, and RNA extraction

NP and nasal swab specimens collected in viral transport medium (VTM) were obtained and processed for qPCR, virus culture, and RNA extraction as previously described (20, 22). We used the same NP swab RNA extracts that had also been used in the qPCR assays for cross-sectional samples. For longitudinal samples (UIUC), we used separate nasal swab RNA extracts since the original extracts used for qPCR were not available. These extracts were from the same VTM as the original extracts and were extracted using the same method.

### SCV-2 gRNA, sgRNA, and RPLP1 ddPCR assays

Since gRNA and all sgRNAs share a common upstream leader sequence (ULS) in the 5’ UTR (Supplemental Figure 1), a previously published common forward (F) primer was used to align within this sequence (5). Four separate reverse (R) primers were designed to amplify either gRNA or the N, E, or S sgRNAs specifically. Although each R primer is capable of binding within the N, E, or S regions that are also found in gRNA or the larger sgRNAs, these longer amplicons (>2000 nucleotides) do not have adequate time to amplify given their massive sizes. Thus, each R primer will specifically only amplify the N, E, or S that is found in their respective sgRNAs. In order to multiplex gRNA assay with either N, E, or S sgRNA assay, a HEX fluorophore was used for the gRNA probe and a FAM fluorophore for sgRNA probes (Supplemental Figure 1; Supplemental Table). We normalized the amount of cellular RNA in each clinical sample RNA extraction using a separate ddPCR to quantify RPLP1 (Ribosomal Protein Lateral Stalk Subunit P1) RNA, an abundant host-derived cellular mRNA, using a commercially available assay (Integrated DNA Technologies; Cat. Hs.PT.58.38347178).

### One-step RT-ddPCR

Clinical sample RNA was added to the components of One-Step RT-ddPCR Advanced Kit for Probes (Bio-Rad; Cat. 1864021) along with appropriate primers and probes, followed by droplet generation and reading according to the manufacturer’s protocol using QX200 Droplet Digital PCR System (Bio-Rad; Cat. 1864001). The PCR conditions for the droplet-partitioned samples were 50⁰C for 60 minutes, 95⁰C for 10 minutes, 40 cycles of 95⁰C for 30 seconds and 57⁰C for 1 minute, 98⁰C for 10 minutes, and then hold at 4⁰C.

### Statistics

For each sample, Poisson statistics were used to calculate the total copy numbers of N sgRNA and gRNA (along with 95% CI) using Bio-Rad QuantaSoft Analysis Pro software. We then converted the copy number to copies/µl based on the VTM volume in which nasal swab specimens were collected. To account for the different efficiencies in RNA extraction and presence of cellular RNA in clinical samples, N sgRNA and gRNA quantities were normalized to RPLP1, which is abundant in nasal epithelial cells. We graphically compared the changes in Ct value, N sgRNA, gRNA, and virus culture results. Kruskal-Wallis tests were used to compare N sgRNA and gRNA quantities between culture-negative and positive samples and to compare Ct values between culture-negative and positive samples. We also calculated sgRNA:gRNA ratios. Spearman correlation coefficients and corresponding p-values were calculated to accommodate data that were not normally distributed.

### Study approval

Informed consent was obtained from the participants in the UIUC cohort. The samples from JHCVL were completely anonymized, and thus could not be tracked to individual participants. This study was approved by the Johns Hopkins University IRB (IRB00254173 and IRB00272523) and Western IRB (20202884).

## Data Availability

All data produced in the present study are available upon reasonable request to the authors.

## Author contributions

HSH designed assays, performed experiments, performed analyses, verified data integrity, and edited the manuscript. CML performed experiments, performed analyses, verified data integrity, and edited the manuscript. MM performed quality control and experiments. TG performed quality control. NG, CHL, MLR, AM, and MC contributed in sample processing and qPCR data generation. AC and RZ contributed in virus culture data generation. CBB provided longitudinal samples and edited the manuscript. AP provided virus culture data and edited the manuscript. HHM and YCM provided samples and qPCR data, and edited the manuscript. CLT and AB conceived of the study, provided funding, performed analyses, and drafted and edited the manuscript. All authors approved the final version of the manuscript.

## Acknowledgements

We would like to thank the participants in this study. This work was supported by the HIV Prevention Trials Network (HPTN) sponsored by the National Institute of Allergy and Infectious Diseases (NIAID), National Heart, Lung and Blood Institute, National Institute on Drug Abuse, National Institute of Mental Health, and Office of AIDS Research, of the NIH, DHHS (UM1 AI068613); R01 AI138810 from NIAID (A.B. and C.T.), U5411090366 (YCM), U54EB007958-13 (YCM, MLR), U54EB007958-12S1 (YCM), and U54HL143541-02S2 (YCM, MLR, NG, CHL, AM, MC, RZ, CBB, HHM) under the NIH RADx-Tech program. The views expressed in this manuscript are those of the authors and do not necessarily represent the views of the National Institute of Biomedical Imaging and Bioengineering; the National Heart, Lung, and Blood Institute; the National Institutes of Health, or the U.S. Department of Health and Human Services. This work was also supported by the JHU COVID-19 Research and Response Program Fund, the Sherrilyn and Ken Fisher Center for Environmental Infectious Diseases Discovery Program, and the Henry Jackson Foundation.

## Supplemental Materials

**Supplemental Figure 1.**
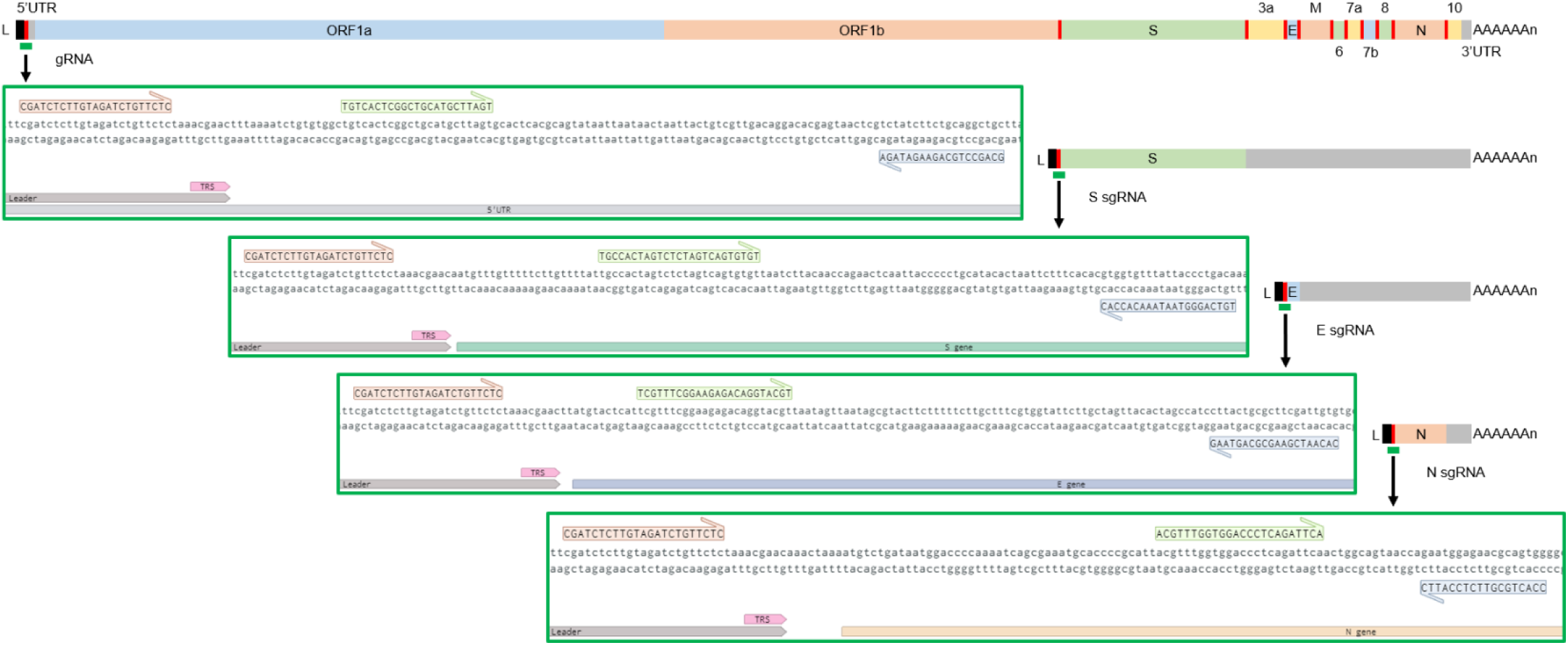
Schematic presentation of SCV-2 gRNA and S, E, and N sgRNA with ddPCR amplicon targets. The black boxes indicate the leader sequence (L) and the red lines indicate the transcription regulatory sequence (TRS) sites. The green bars indicate ddPCR amplicon targets with magnified view in green box showing primer and probe binding sites (the common leader forward primers in orange, target specific probes in green, and target specific reverse primers in blue). The complete genome sequence of Wuhan-Hu-1 SCV-2 isolate (NCBI GenBank NC_045512.2) was used as a reference sequence.

**Supplemental Table.**
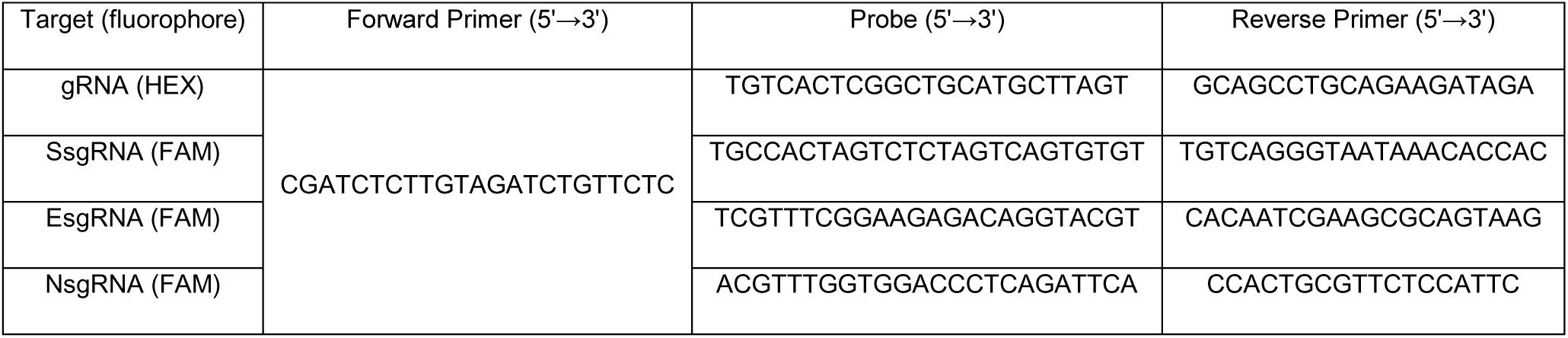
List of ddPCR primers and probes with sequences.

**Supplemental Figure 2.**
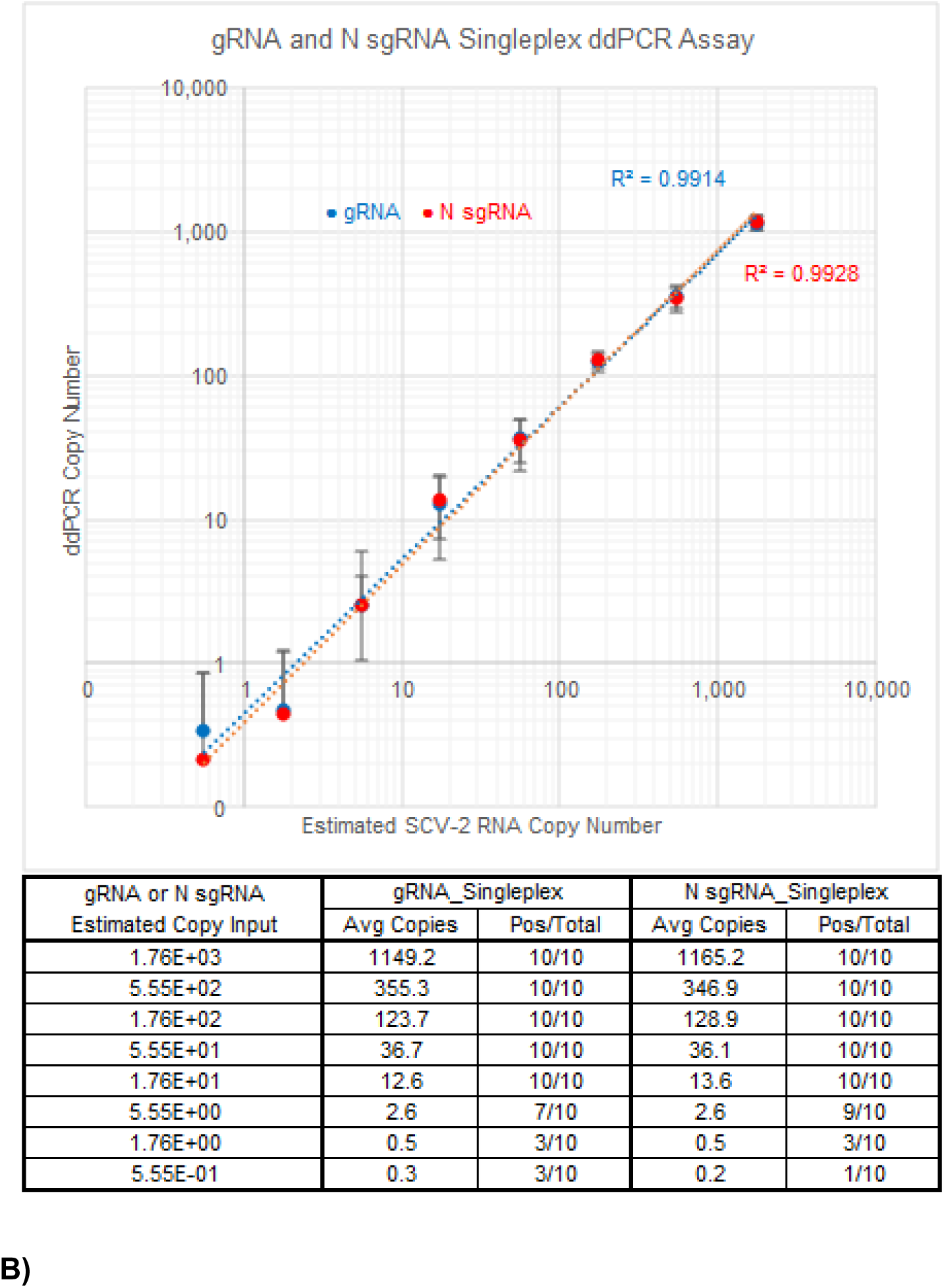
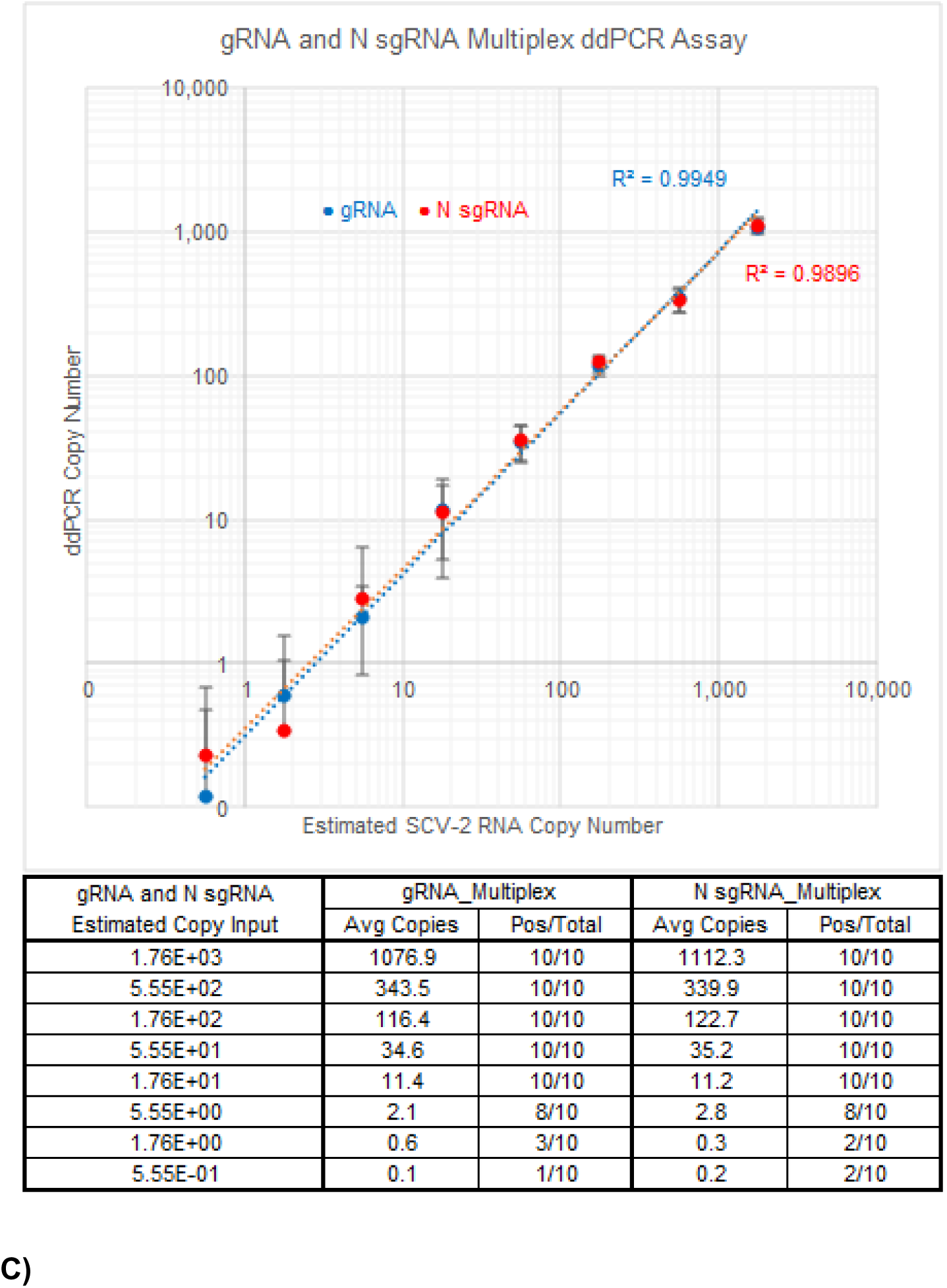
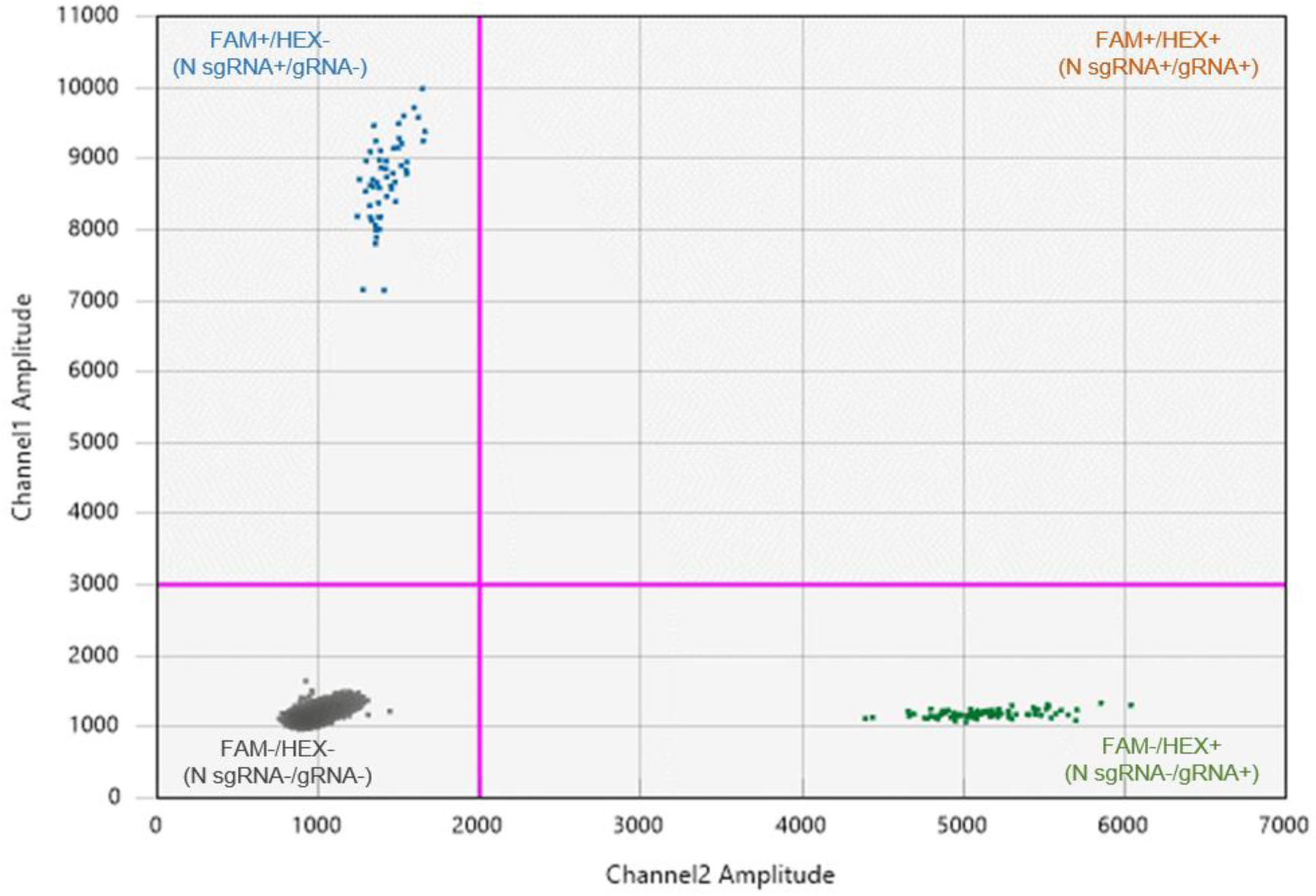
ddPCR sensitivity and specificity for sgRNA and gRNA individually and in multiplex. Synthetic DNA fragments including the T7 promoter sequence and SCV-2 gene fragments for each of the target N, E, S sgRNAs and gRNA were in vitro transcribed to produce each targeted RNA fragment. Half-log dilutions of each RNA ranging from 1755 to 0.56 copies were used to test the performance of sgRNA and gRNA ddPCR assays for each target in A) singleplex and B) multiplex assays. All gRNA and N, E, and S sgRNA assays had similar efficiencies, but N sgRNA assay was integrated into a multiplexed assay with gRNA assay in the subsequent testing because N sgRNA is more abundant in nasal swabs than E and S sgRNAs (data not shown). N sgRNA and gRNA assays were performed A) individually and B) in multiplex on the corresponding RNA dilutions in ten replicates. The tables below each figure indicate the expected versus the observed number of copies of each target, and the number of replicates that were positive at each dilution. Error bars indicate standard deviation. C) The ddPCR plot for a representative sgRNA and gRNA multiplex assay is shown, demonstrating the specificity of each assay by the exclusion of detection of each target by the opposite assay. The probe for gRNA is HEX+ and for N sgRNA is FAM+. Each droplet contains at most one template (gRNA or sgRNA). If a droplet contains sgRNA, then it would register as FAM+/HEX- (blue cluster of points), and if it contains gRNA then it would register as FAM-/HEX+ (green cluster). If there was no SCV-2 template, then it would register as negative for both probes (black cluster). False detection of an sgRNA as a gRNA, or vice versa, would result in a FAM+/HEX+ cluster, which is not observed here.

**Supplemental Figure 3.**
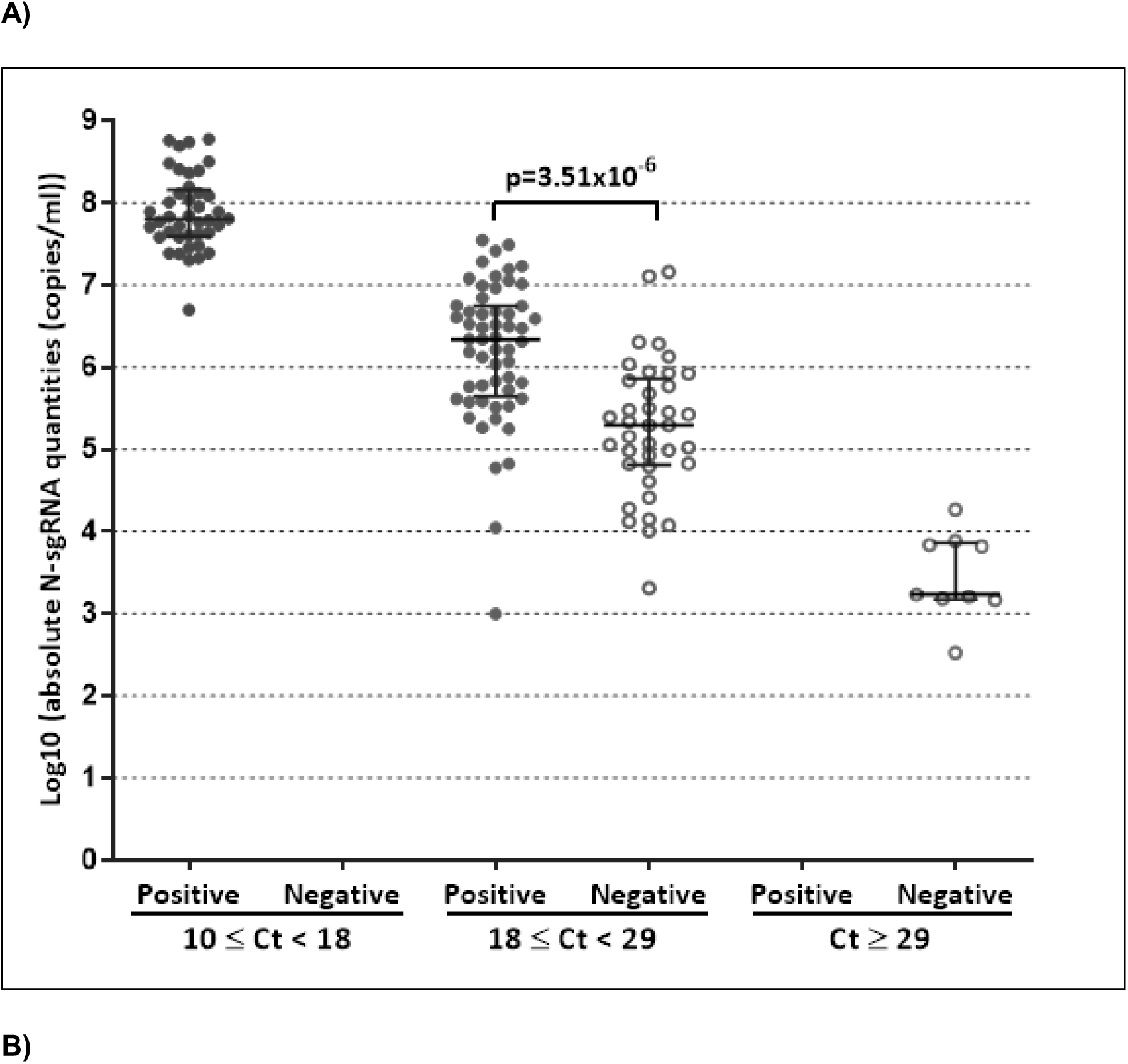
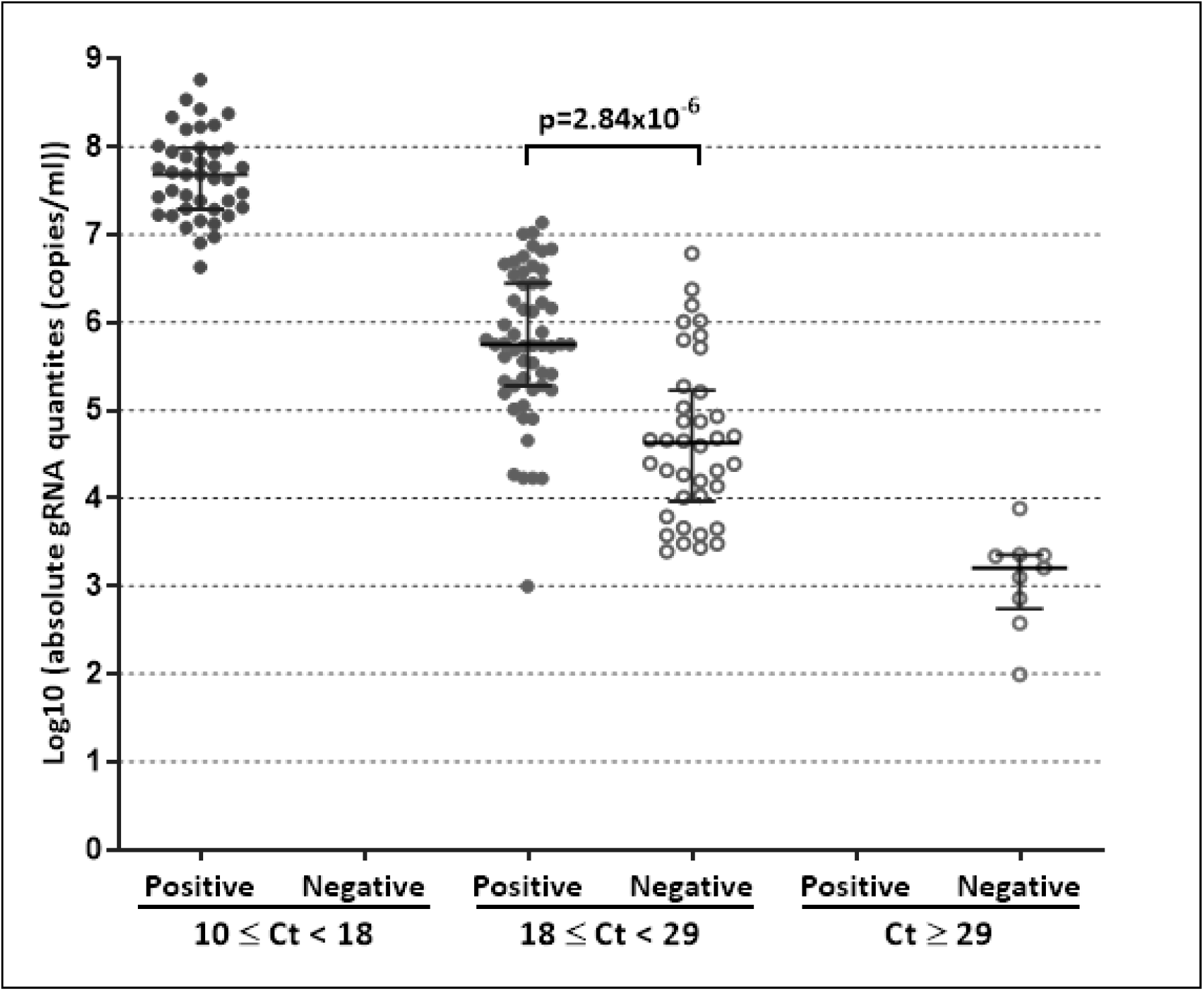
Absolute N sgRNA and gRNA quantities, total SCV-2 RNA quantities, and virus culture results. Shown are the absolute quantities of A) N sgRNA and B) gRNA, stratified by total SCV-2 RNA quantities as measured by qPCR and virus culture results. P-values calculated by Kruskall-Wallis tests are shown.

**Supplemental Figure 4.**
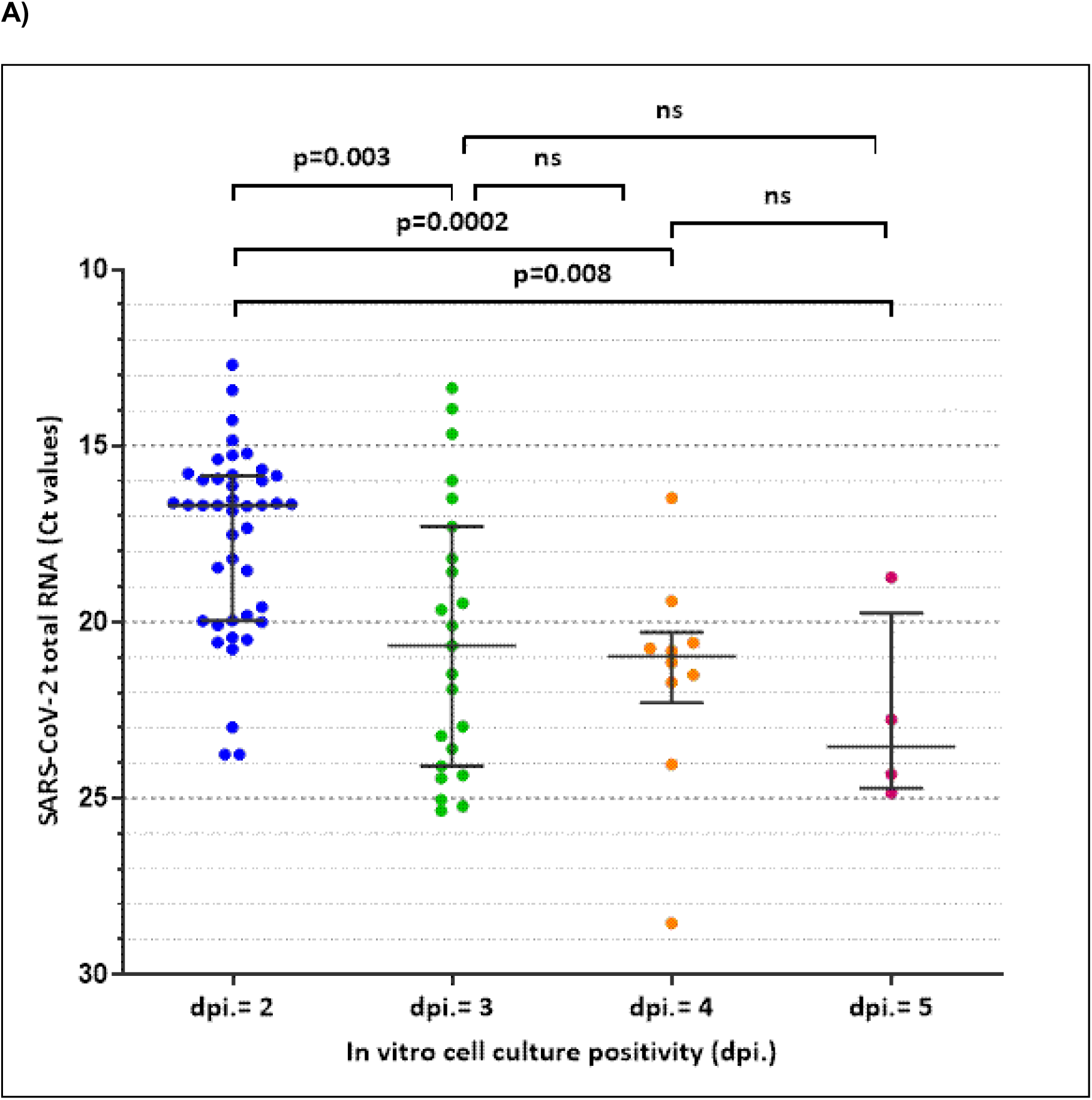
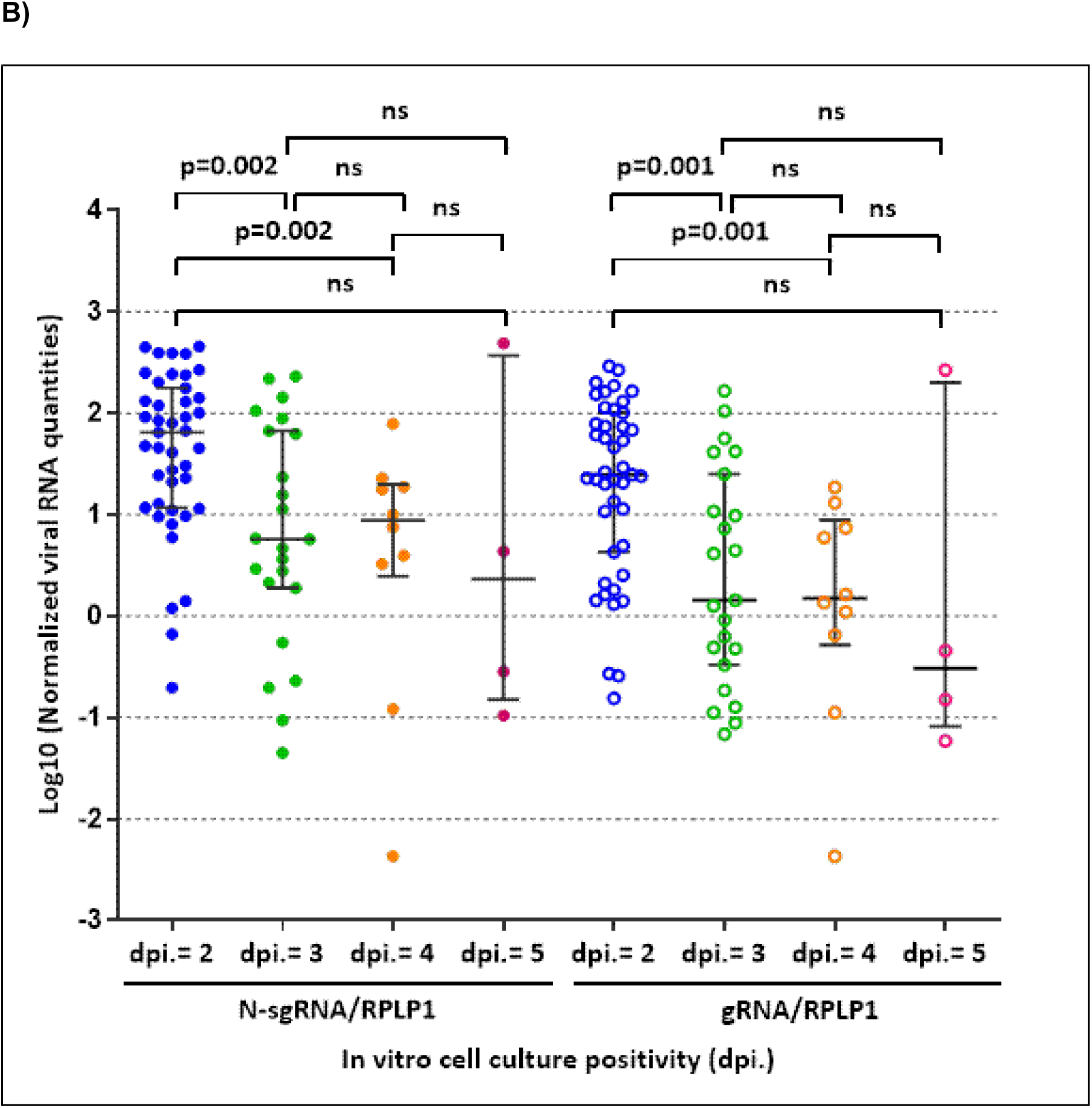
N sgRNA, gRNA, and total SCV-2 RNA amounts correspond with the rapidity of culture positivity. Aliquots from the same NP swabs obtained from 144 JHCVL samples were tested for N sgRNA and gRNA (by multiplex ddPCR), host gene RPLP1 (by singleplex ddPCR), total SCV-2 RNA (by qPCR), and virus culture. Virus culture is reported as positive or negative but is also annotated for the number of days post infection (dpi) that it turns positive. A low dpi is indicative of abundant infectious virions while a high dpi is indicative of fewer infectious virions. Thus, dpi can be used as a separate semi-quantitative measurement of virus quantities. A) N sgRNA and gRNA quantities, adjusted for RPLP1 levels, and B) total SCV-2 RNA quantities are shown for all culture-positive samples, stratified by dpi. P-values, calculated by Kruskal-Wallis test, are shown for each comparison.

**Supplemental Figure 5.**
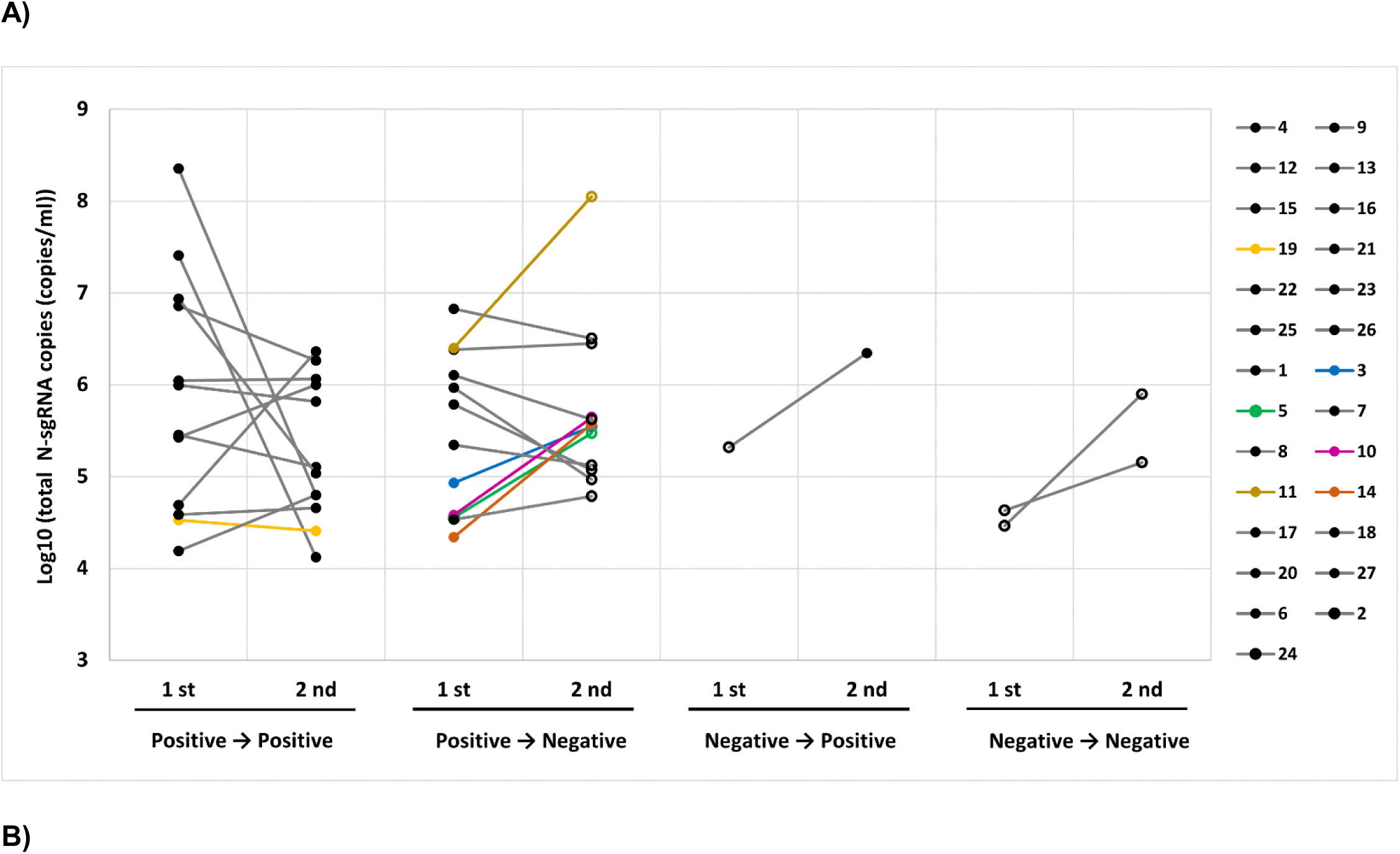
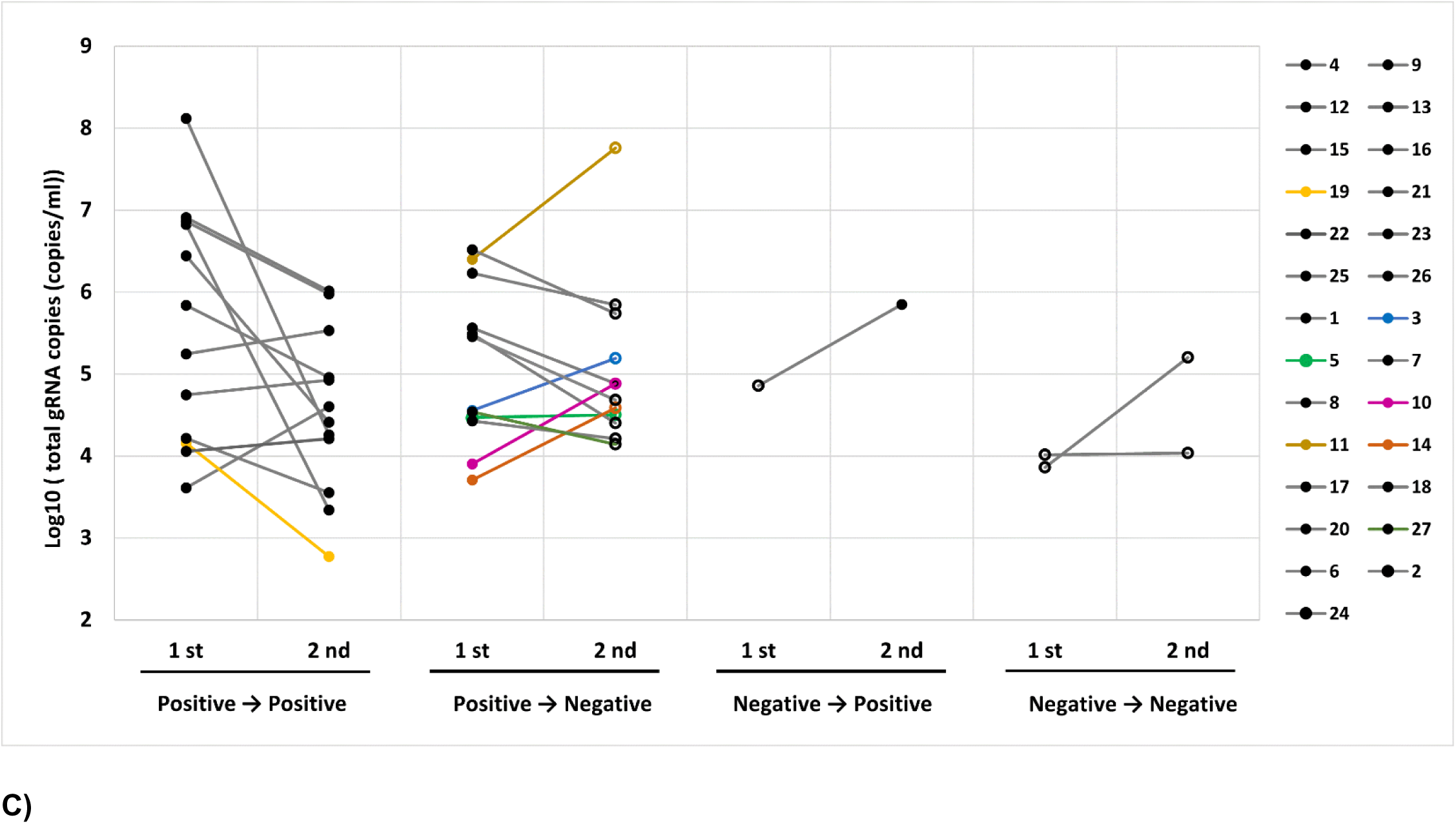
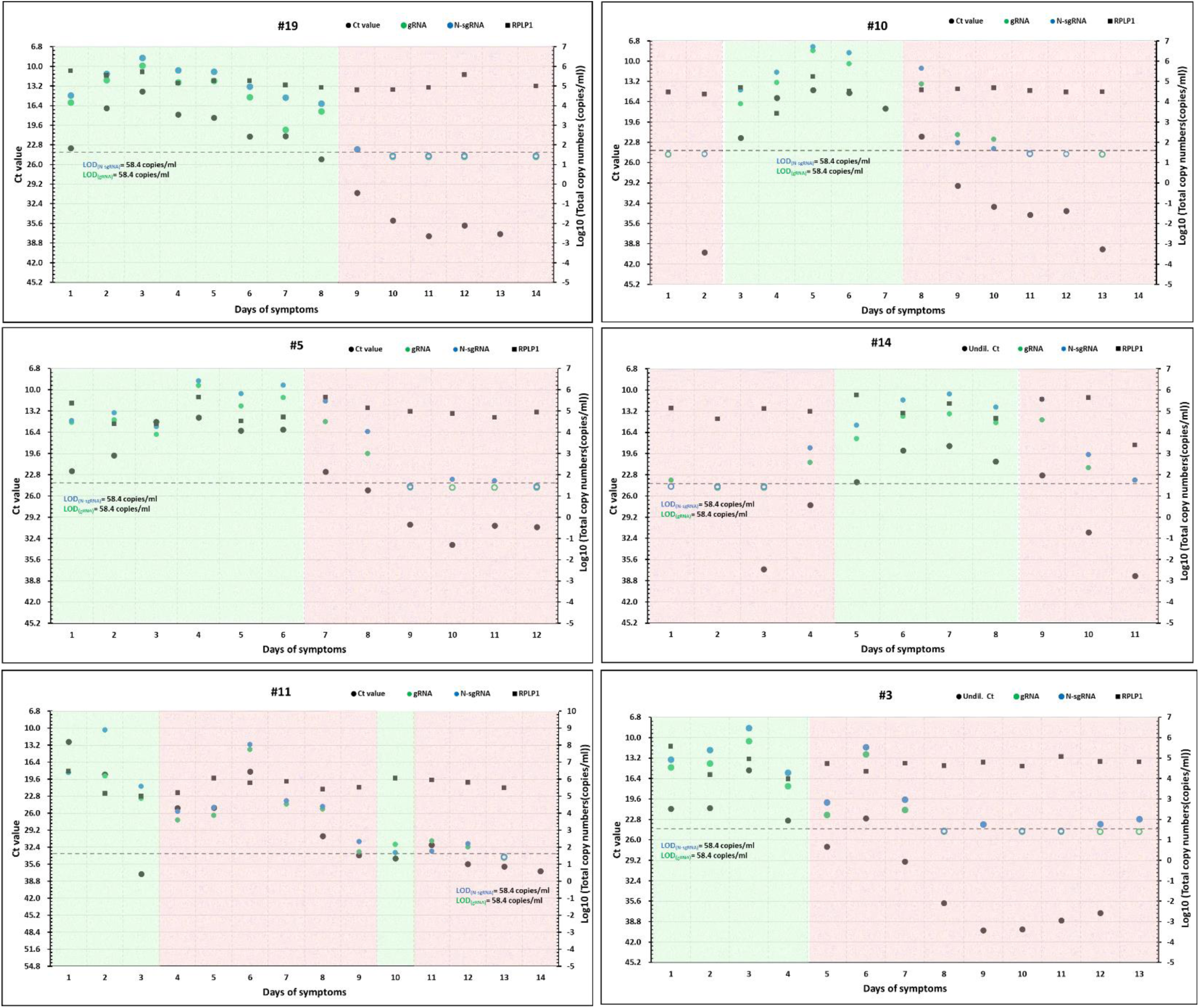
Absolute daily quantities of N sgRNA, gRNA, total SCV-2 RNA, and culture results. In 27 persons from the UIUC cohort with paired samples available, absolute amounts of A) N sgRNA and B) gRNA are shown in early and late time points after symptom onset. The 27 persons were categorized into four groups based on virus culture results from contemporaneous samples: those that were culture-positive early and late (Pos→Pos), those that were culture-positive early and culture-negative late (Pos→Neg), those that were culture-negative early and culture-positive late (Neg→Pos), and those that were culture-negative at both times (Neg→Neg). Legend and colors indicate participant ID number. C) In 6 persons in the UIUC cohort, daily NP swabs were tested for multiple RNAs: N sgRNA (by ddPCR), gRNA (by ddPCR), total SCV-2 RNA (by qPCR), and RPLP1 (by ddPCR). Virus culture was also performed on daily samples. Shown are total SCV-2 RNA (Ct values – black points), absolute log_10_ N sgRNA values (blue points), absolute log_10_ gRNA values (green points), RPLP1 (black squares) and virus culture results (pink shading – culture-negative; green shading – culture-positive). Lower limits of detection for N sgRNA and gRNA are shown as horizontal hashed lines.

**Supplemental Figure 6.**
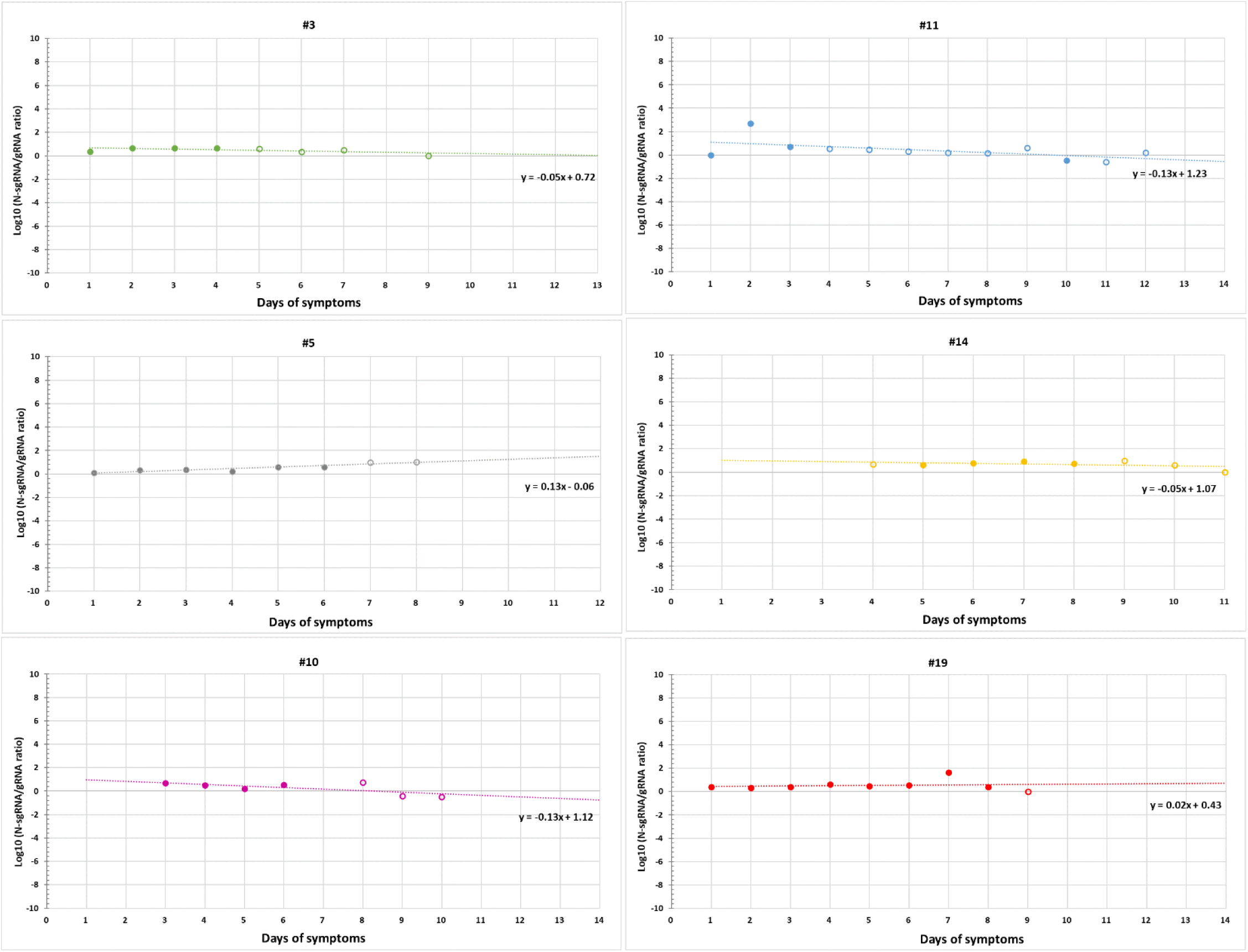
Individual trajectories of N sgRNA:gRNA ratios from daily samples. Each of the six individuals with daily N sgRNA and gRNA results had N sgRNA:gRNA ratios calculated for each sample. The ratios were plotted for each day to identify a trajectory of ratios, as illustrated in Figure 6. Shown is each individual trajectory.

## Notes

**Conflict of interest:** YCM has received research grant support to Johns Hopkins University from Hologic, Ceres, Cepheid, Roche, ChemBio, Becton Dickinson, miDiagnostics, and Yukon, and has provided consultative support to Abbott. The other authors have declared that no conflict of interest exists.

### Competing Interest Statement

YCM has received research grant support to Johns Hopkins University from Hologic, Ceres, Cepheid, Roche, ChemBio, Becton Dickinson, miDiagnostics, and Yukon, and has provided consultative support to Abbott. The other authors have declared that no conflict of interest exists.

